# Integrating dynamical modeling and phylogeographic inference to characterize global influenza circulation

**DOI:** 10.1101/2024.03.14.24303719

**Authors:** Francesco Parino, Emanuele Gustani-Buss, Trevor Bedford, Marc A. Suchard, Nídia Sequeira Trovão, Andrew Rambaut, Vittoria Colizza, Chiara Poletto, Philippe Lemey

**Affiliations:** Sorbonne Université, INSERM, Institut Pierre Louis d’Epidemiologie et de Santé Publique (IPLESP), Paris, France; Department of Microbiology, Immunology and Transplantation, Rega Institute, KU Leuven – University of Leuven, 3000 Leuven, Belgium; Vaccine and Infectious Disease Division, Fred Hutchinson Cancer Center, Seattle, Washington 98109, USA; Howard Hughes Medical Institute, Seattle, Washington 98109, USA; Departments of Biomathematics and Human Genetics, David Geffen School of Medicine at UCLA, University of California, Los Angeles, CA, 90095, USA; Department of Biostatistics, UCLA Fielding School of Public Health, University of California, Los Angeles, CA, 90095, USA; Fogarty International Center, National Institutes of Health, Bethesda, MD, USA; Institute of Ecology and Evolution, University of Edinburgh, Edinburgh EH9 3FL, UK; Department of Biology, Georgetown University, Washington, DC, USA; Department of Molecular Medicine, University of Padova, 35121 Padova, Italy

**Author notes:** F.P. and E.G.-B. contributed equally to this work. V.C., C.P. and P.L. contributed equally to this work.

**Keywords:** Influenza, Metapopulation, Phylogeography, Bayesian inference

## Abstract

Global seasonal influenza circulation involves a complex interplay between local (seasonality, demography, host immunity) and global factors (international mobility) shaping recurrent epidemic patterns. No studies so far have reconciled the two spatial levels, evaluating the coupling between national epidemics, considering heterogeneous coverage of epidemiological and virological data, integrating different data sources. We propose a novel combined approach based on a dynamical model of global influenza spread (GLEAM), integrating high-resolution demographic and mobility data, and a generalized linear model of phylogeographic diffusion that accounts for time-varying migration rates. Seasonal migration fluxes across global macro-regions simulated with GLEAM are tested as phylogeographic predictors to provide model validation and calibration based on genetic data. Seasonal fluxes obtained with a specific transmissibility peak time and recurrent travel outperformed the raw air-transportation predictor, previously considered as optimal indicator of global influenza migration. Influenza A subtypes supported autumn-winter reproductive number as high as 2.25 and an average immunity duration of 2 years. Similar dynamics were preferred by influenza B lineages, with a lower autumn-winter reproductive number. Comparing simulated epidemic profiles against FluNet data offered comparatively limited resolution power. The multiscale approach enables model selection yielding a novel computational framework for describing global influenza dynamics at different scales - local transmission and national epidemics vs. international coupling through mobility and imported cases. Our findings have important implications to improve preparedness against seasonal influenza epidemics. The approach can be generalized to other epidemic contexts, such as emerging disease outbreaks to improve the flexibility and predictive power of modeling.

## 1 Introduction

Seasonal influenza viruses cause recurrent epidemics characterized by annual periodicity in temperate countries and by diverse, less regular patterns in the tropics.^1^ Extensive epidemiological research highlighted the critical role of local sociodemographic and environmental aspects (e.g. weather conditions, school calendar and increased indoor activity) in the onset and unfolding of influenza waves.^2–10^ At the same time, international human travel ensures rapid worldwide circulation of influenza .^11–13^ Phylogeographic studies have reconstructed the global migration patterns of seasonal influenza in extensive detail, revealing limited local persistence of the virus in most regions and highlighting the importance of continual reseeding in determining viral genetic structure, severity and timing of the epidemics.^12,14–16^ The concurrent impact of local and global drivers has also been apparent through analyses of the decline of influenza incidence observed during the coronavirus disease 2019 (COVID-19) pandemic. Both reduction in international travel and social restrictions (due, e.g., to remote working and school closure) were, indeed, found to be associated to the influenza drop.^17–19^ From a modeling perspective, however, reconciling the two spatial levels represents a major challenge, as the interplay between the local progression of an epidemic in a particular country and the coupling between different epidemics mediated by human mobility remains poorly understood.

The unevenly distributed human host population across countries and seasonal areas and the complex network of human travel acting over both short and long range distances (i.e. from city-to-city commuting to international air travel) are key to this challenge.^20,21^ In addition, while influenza surveillance is improving worldwide,^18^ the coverage is still biased, restricting our ability to resolve the spatial dynamics. The quality of epidemiological data is generally higher in temperate areas, but these are the areas that are characterized by a high degree of synchronization of national influenza epidemics in the same hemisphere, thus making spatial effects less identifiable. As a consequence, mathematical models for influenza dynamics at different scales remain difficult to parametrize. Genetic data however carry the signature of large-scale circulation dynamics and may therefore represent a valuable complementary source to characterize seasonal influenza epidemics, especially when combined with epidemiological data. The recent severe acute respiratory syndrome coronavirus 2 (SARS-CoV-2) pandemic has illustrated the importance of combining epidemiological and mobility data, genomic sequences and metadata to provide insights into viral emergence and spread.^22–27^ Phylogeographic inference in particular has been widely applied to elucidate SARS-CoV-2 genomic epidemiology, including origins, introductions, routes of dispersal, and drivers associated with variant dissemination, contributing to the effectiveness of systematic genomic surveillance.^24–27^ However, the development of integrated tools is still relatively limited and opportunities remain to fulfill the full potential of phylodynamic approaches. Different from previous phylogeographic reconstructions, we here propose a novel approach that combines a high-resolution dynamical model for the diffusion of influenza worldwide, informed by extensive demographic and mobility data, and a generalized linear model of phylogeographic diffusion that allows for inhomogeneous migration rates over time. We use the epidemic model to simulate migration fluxes across macro regions of the world and evaluate their ability to explain phylogeographic patterns.

## 2 Results and Discussion

### 2.1 Combining mechanistic epidemic modeling and phylogeographic inference

We combine dynamical modeling and phylogeographic inference by considering the simulated fluxes of infectious cases generated by a data-driven computational model for infectious disease spread at the global scale as predictors for phylogeographic migration rates. For this purpose, we build on GLEAM, the GLobal Epidemic and Mobility model,^28^ that integrates high-resolution demographic and mobility data at different spatial scales - air traffic database comprising nearly all commercial air-travels and short-range mobility obtained from national commuting data. ^28^ The global population is distributed among 3362 patches corresponding to large urban areas and traveling of individuals is modeled explicitly based on passenger data. GLEAM has been used to model the short-term outbreak dynamics in the case of the H1N1 influenza pandemic,^11,13^ Ebola, ^29,30^ MERS,^31,32^ Zika, ^33^ and COVID-19, ^34–37^ following prior modeling work considering the global scale.^38–40^

To adapt GLEAM to seasonal circulation of influenza, we introduce a more realistic scheme for modeling mobility of individuals that preserves their geographic residence (see Material and Methods for more details). This approach is usually adopted for modeling recurrent travel, such as commuting, ^28,41,42^ while a simpler Markovian mobility model assuming memory-less traveling trajectories is generally preferred for air travel. Although less realistic, the latter is more parsimonious and has only a limited approximation bias for fast spreading diseases and short-term epidemic dynamics.^42^ In our analysis of the multi-annual influenza propagation, we compare both the Markovian and the recurrent travel approach.

In the metapopulation scheme, GLEAM transmission dynamics occur within patches ruled by a compartmental model specific to seasonal influenza^2,3,11,43,44^ (Material and Methods) and accounting for: (i) a temporary immunity to the virus of average duration *D*, with values explored between 1 and 8 years; (ii) a geographically dependent seasonal transmission in temperate areas varying sinusoidally in time between a minimum and a maximum basic reproductive number, *R*_min_ (explored values: 0.5 and 0.75) and *R*max 2 [1.25,2.5], respectively, with 15 Nov, 15 Dec, and 15 Jan tested as dates of maximum transmission in the northern hemisphere and minimum transmission in the southern hemisphere; (iii) a constant transmission with a basic reproductive number equal to *R*max in the tropics. Discrete stochastic simulations at the individual level provide numerical trajectories for the global seasonal dynamics with a time resolution of one day. Results display autumn-winter waves interspersed by subcritical spring-summer transmission in the temperate hemispheres, and an irregular continuous circulation in the tropics. The output is summarized as fluxes during the spring-summer and autumn-winter epochs between the countries considered by the phylogeographic approach.

To evaluate different parameterisations of our dynamical model, we test the resulting fluxes in a phylogeographic approach. To this purpose, we adopt a generalized linear model (GLM) extension of discrete phylogeographic diffusion ^12^ that accommodates time-inhomogeneous migration dynamics. The GLM-diffusion approach allows modeling the intensity of location exchange between discrete states along a phylogeny as a function of a number of potential predictors.^12^ Using epoch modeling,^45^ we allow for different location exchange processes, and hence different predictors, across different time intervals in the evolutionary history. Specifically, we consider the difference in model-based fluxes during alternating spring-summer and autumn-winter epochs. We employ comprehensive sequence data sets that were previously analyzed to reconstruct more than a decade of global seasonal migration dynamics of influenza A H3N2, H1N1 (prior to the H1N1/09 pandemic), B Victoria (VIC) and Yamagata (YAM) between 2000 and 2012. ^16^ We model the phylogeographic process between the countries of sampling for these different influenza subtypes. We use BEAST^46^ to estimate parameters of the time-inhomogeneous GLM-diffusion approach through Bayesian inference while averaging over a set of time-measured trees.

### 2.2 Model selection and parameter estimation

We evaluated different sets of migration fluxes, including simple passenger fluxes based on air travel data, simulated migration fluxes from the Markovian and the recurrent travel version of GLEAM, either as homogeneously aggregated fluxes over time (annual fluxes) or as two-epoch level fluxes (seasonal fluxes).

To avoid including an excessive amount of migration flux predictors in a single GLM diffusion model analysis, our systematic evaluation relied on a stepwise approach. We first tested Markovian against recurrent travel fluxes for different epidemiological parameters (*R*_min_, *R*max and *D*) while conditioning on seasonal fluxes with a peak time in January in the northern hemisphere. Next, we tested seasonal against annual fluxes conditioning on recurrent travel fluxes and a peak time in January. Finally, we tested the January peak time against peak times in December and November conditioning on seasonally aggregated recurrent travel fluxes. In each of these analyses, we include simple air passenger fluxes as a baseline predictor, and we perform the analysis with and without a predictor based on the residuals of a regression of sample sizes against population sizes to assess the potential impact of sample sizes. Fig. 1 summarizes the marginal posterior inclusion probabilities for the different comparisons, while inclusion probabilities for individual flux predictors are provided in Supplementary Tables 3-6.

**Figure 1:**
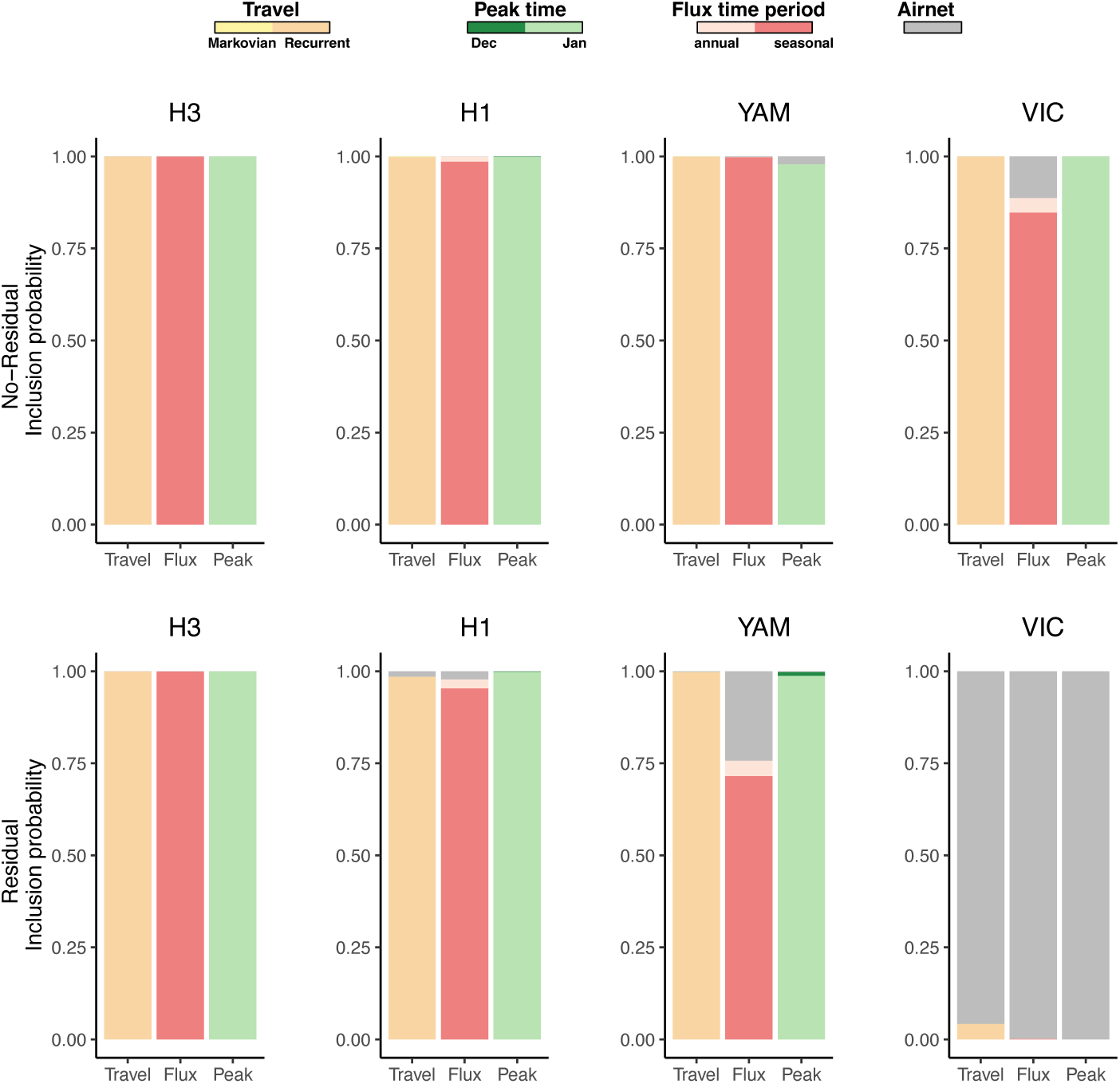
Marginal posterior inclusion probabilities associated with air travel data or with GLEAM-based fluxes comparing recurrent against Markovian travel, seasonal against annual fluxes and different peak time times. For peak times, we performed an analysis comparing Nov 15 against Dec 15 and an analysis comparing Dec 15 to Jan 15, but we only show the latter for simplicity as Dec 15 outperformed Nov 15.

Our analyses support GLEAM fluxes based on recurrent travel for H3N2, H1N1 and YAM, demonstrating i) an improvement of dynamical model predictions over simple air travel for most of the influenza variants and ii) the importance of accounting for memory in the origin of travel trajectories of individuals. For these three variants, seasonally aggregated fluxes also outperformed annual fluxes and a peak time in January outperformed earlier peak times (with an inclusion probability of ⇠1). For H3N2 specifically, seasonal fluxes strongly outperformed annual fluxes with and without residual predictor (inclusion probability of ⇠1) (Supplementary Table 4) and a similar support was detected for H1N1 (inclusion probability of ⇠0.95). The inclusion probability for YAM was marginally lower when employing the residual predictor (0.72). When comparing the magnitude of best-supported predictors, the support for seasonal fluxes was approximately 20 times stronger than annual flux. In the case of VIC, GLEAM fluxes are only supported in the analysis without sample size residual. Standard phylogeographic analyses of these data sets have previously shown that VIC is associated with the highest degree of persistence and the lowest overall migration,^16^ so offering less information to support GLEAM-based fluxes, in particular when sample heterogeneity can explain a considerable degree of migration variability. For the seasonal GLEAM fluxes based on recurrent travel and based on a Jan 15 peak time, we next summarized the support for the different values of *R*max, *R*_min_ and *D* (Fig. 2). For H3N2, the parameter combination including *R*max = 2.25, *R*_min_ = 0.75 and *D* = 2 years yields the highest flux inclusion probability (inclusion probability ⇠1 and 0.68 with and without sample size residual, respectively). The parameter values in this combination are also clearly the ones that are preferred across all combinations in analyses with and without residual predictor (Fig. 2). In Section 3 of the Supplementary Material, we investigate differences in fluxes generated by different parametrizations and note that the GLM selects fluxes that are distinct from those generated by other parameterizations. The H1N1 analysis with sample size residual also finds this parameter combination to be the best supported, but not as strongly so (0.59 inclusion probability), while the analysis without sample size residual finds marginally lower support (0.33 inclusion probability) compared to a parameter combination including *R*max = 1.50, *R*_min_ = 0.75 and *D* = 1 years (0.52 inclusion probability).

**Figure 2:**
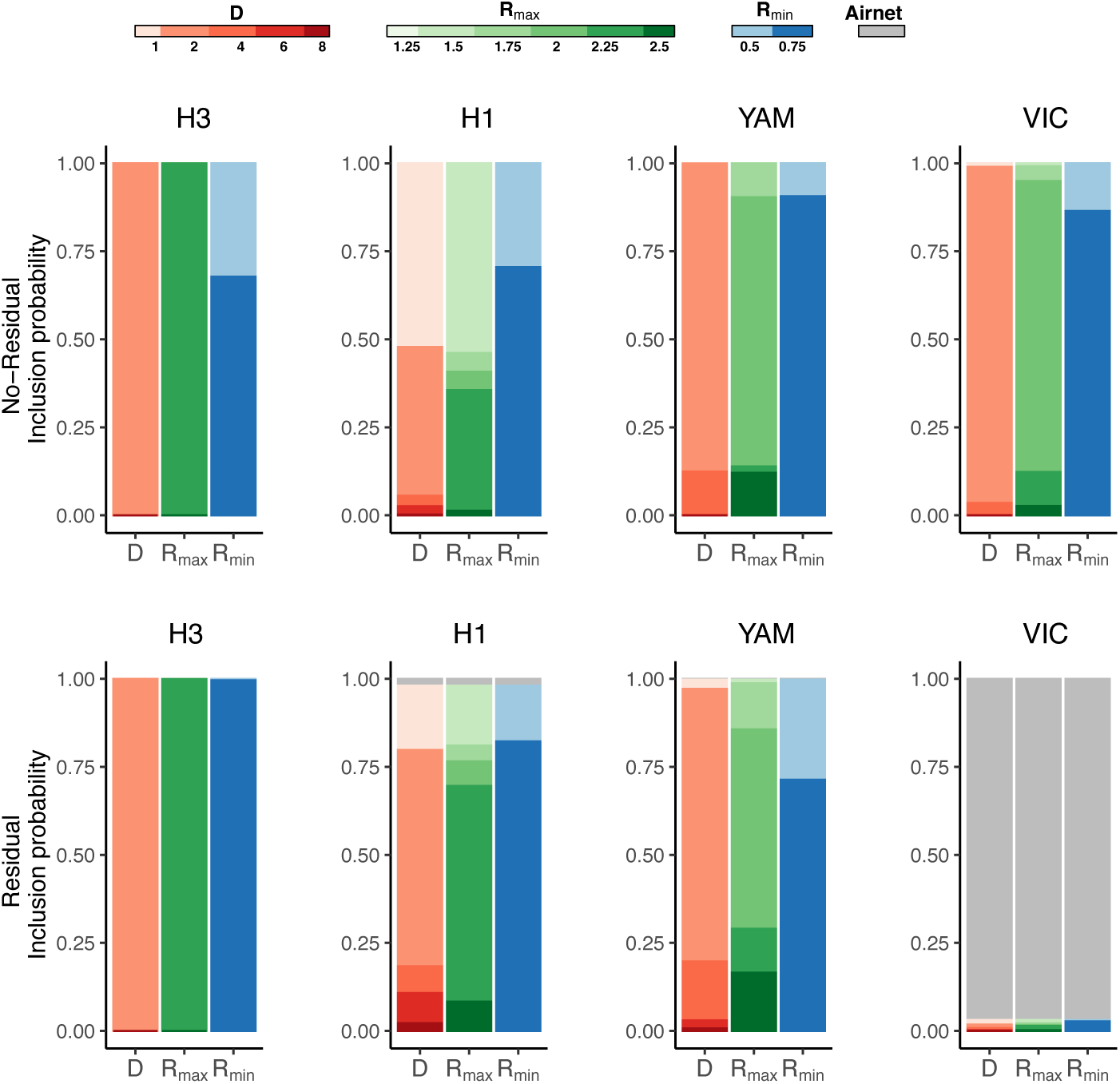
Marginal posterior inclusion probabilities associated with recurrent travel distribution of GLEAM fluxes for the three parameters *R*max, *R*_min_ and *D*, without and with residual predictor.

The analysis of YAM without residual predictor recovers the same general preference for *D* and *R*_min_ (2 years and 0.75, respectively), but finds the strongest support for a somewhat lower *R*max of 2.00 (0.77 inclusion probability). The same parameterization is supported by the analysis with residual predictor, but with somewhat lower inclusion probability (0.56). Without residual predictor, the analysis of VIC supports the same parameterization as YAM (0.60 inclusion probability, Fig. 2).

The rapid immune waning we identify, with average duration of immune protection between 1 and 3 years, is in agreement with previous work,^47–50^ but some studies have estimated longer immunity duration.^2,3^ Smaller transmissibility for influenza B compared to A/H3N2 (as estimated for both Yamagata and Victoria) is also consistent with previous works.^16,48^ Those studies typically use incidence curves at the country level, whereas our selection is based on influenza migration patterns encoded in the genetic data. The spatial coupling, thus, carries the signature of country seasonal waves. For the period between two consecutive epidemics, early analyses of genetic data in temperate areas suggested that inter-seasonal circulation of influenza was dominated by the importation of cases from other seasonal areas, with negligible local transmission following importation.^51–53^ Recent improvements in out-of-season surveillance are providing increasing evidence for the sporadic generation of cases during the spring-summer period,^54–56^ in agreement with spring-summer transmissibility of *R*_min_ = 0.75 supported by our analyses.

### 2.3 Predicted influenza dynamics

We compare country-level epidemics simulated by GLEAM with recurrent travel and epidemic profiles reconstructed from FluNet data to evaluate model predictions against available surveillance data. The number of influenza-positive samples stored for each subtype in FluNet has been extensively used for reconstructing the timing and shape of the epidemic peaks.^1,8,17,47,57^ We compute monthly distributions of cases averaged over the period 2004/05—2014/15 as an indicator of the typical influenza behavior of the country (see Supplementary Material for additional details). Fig. 3 compares the H3N2 epidemic profile with the simulated one for a set of countries in each seasonal area (see Supplementary Material for additional details). The timing and shape of the epidemic waves are well reproduced by the model in the majority of temperate area countries (average correlation 0.83 ± 0.03 for the Northern hemisphere and 0.66 ± 0.10 for the Southern hemisphere), representing also the region with the highest availability of country records. In the tropics, correlations are generally lower (average correlation 0.10 ± 0.05) because of the rather noisy and flat epidemic profiles. For large countries, strong spatial fragmentation of the population and climatic heterogeneity complicates analyses at the national scale,^58^ as illustrated for China. These results were obtained with the best parameterization for H3N2 as selected by the phylogeographic GLM. Other parameter sets performed worse, but a number of different simulated scenarios also showed a similarly high correlation with the data (see Supplementary Material for details and analysis of other influenza subtypes). These results indicate that the degree of information carried by incidence data is limited, likely due to the high level of synchronization between country waves, together with non-uniform surveillance coverage and quality (see Supplementary Fig. 5). This suggests that the best parametrization could be selected as the one that has high correlation with incidence and at the same time explains the genetic data well, i.e. has high inclusion probability in the GLM analysis.

**Figure 3:**
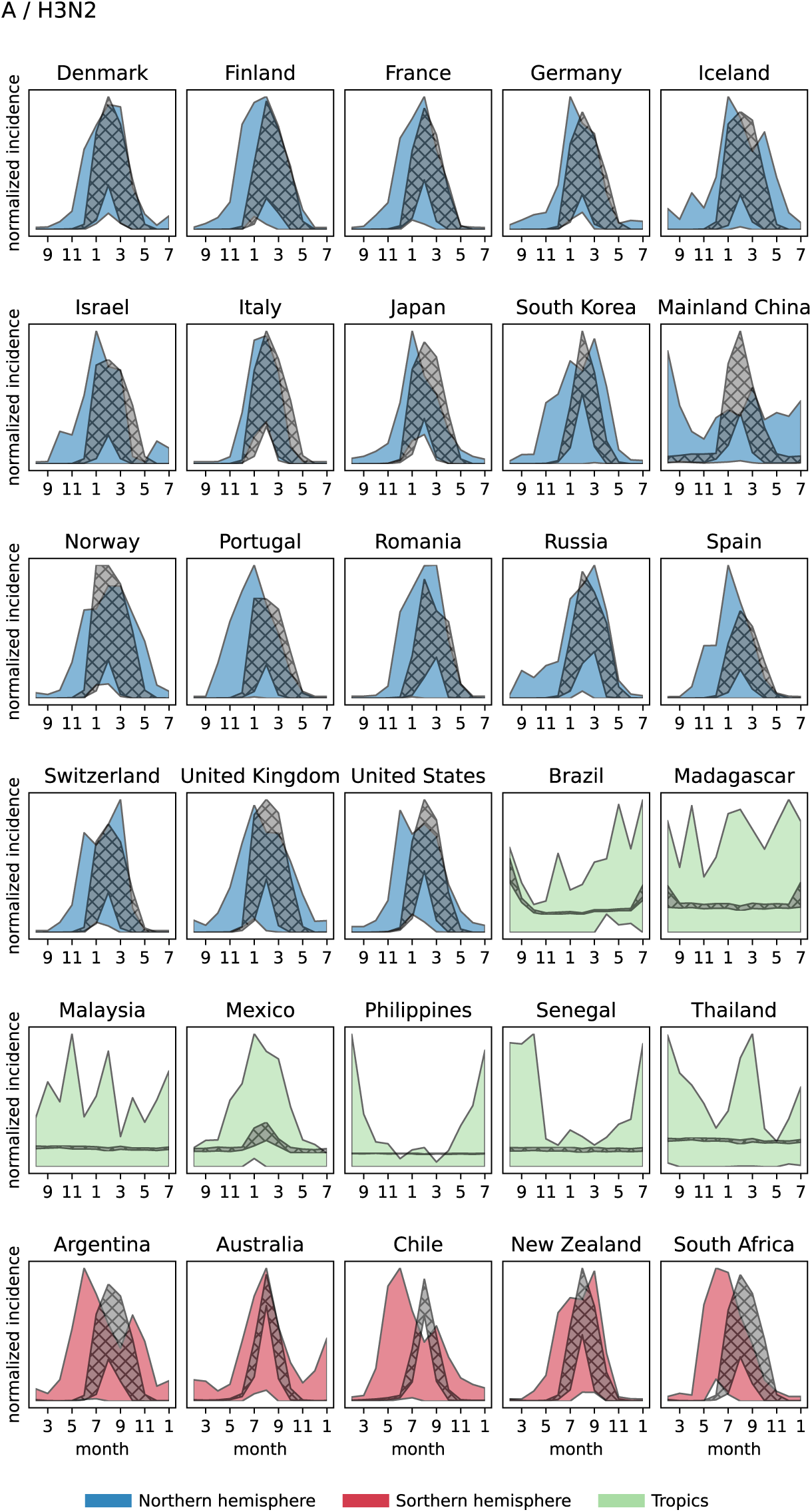
Annual epidemic profiles for 30 selected countries from FluNet H3N2 samples (colored) and simulations with the best-supported scenario (gray). For each seasonal region, selected countries are representative of the whole set for average correlation and its dispersion. Shaded areas show the 95% CI of the normalized incidence (see Supplementary Material).

Simulated influenza circulation shows a strong coupling between Europe and North America during autumn-winter, together with the central role of Southeast Asia and China as influenza sources for the Asian continent and Oceania during spring-summer and autumn-winter periods. In particular, Southeast Asia is one of the main sources of importation for Australia, Japan and Korea, India and Europe. South America appears to be disconnected from Asian regions (see Fig. 4 and Supplementary Table 8 and 9), but plays a role as seeder of influenza in North America and Europe during spring-summer epochs.

**Figure 4:**
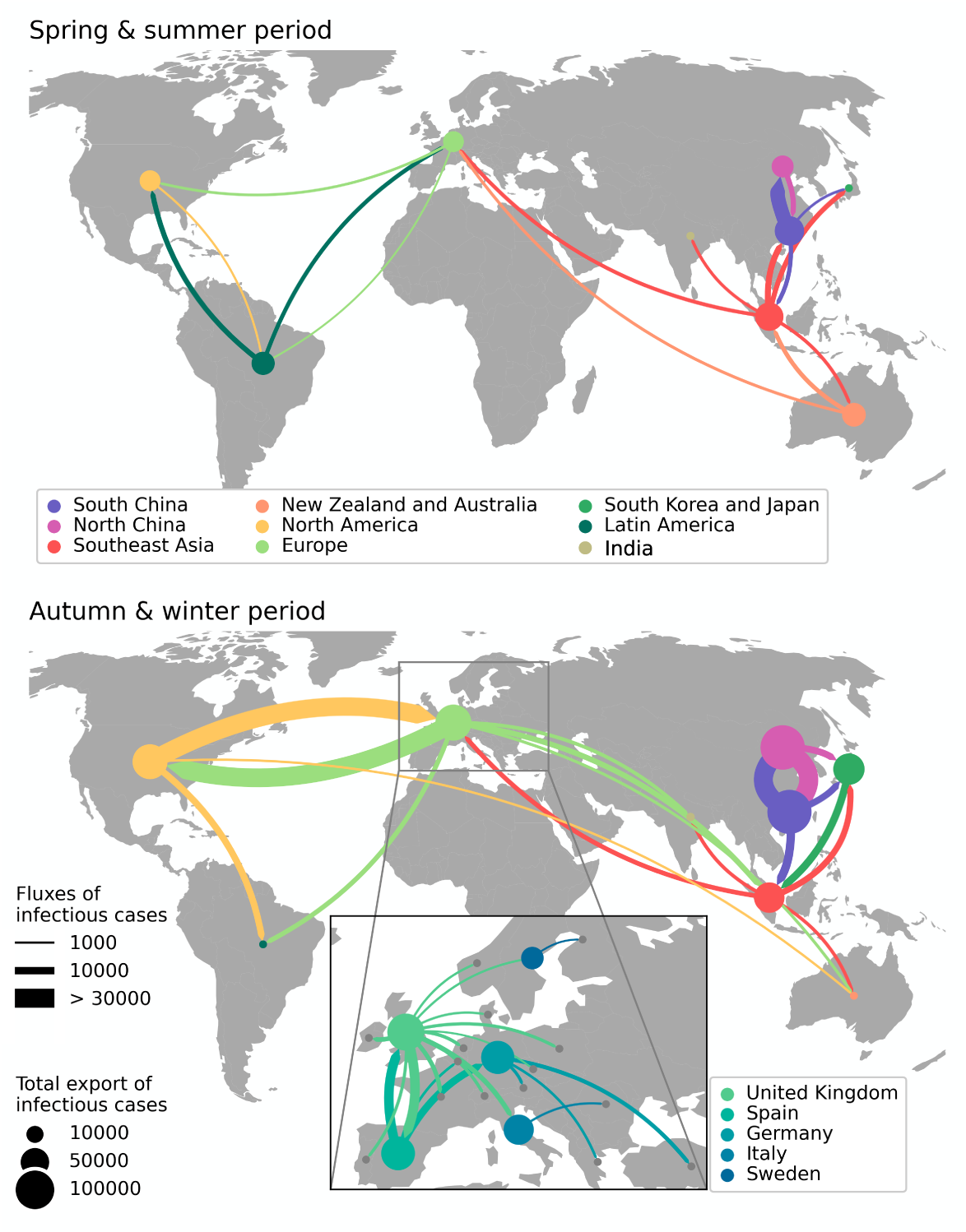
Dominant fluxes of cases for the two epochs. To enhance clarity, countries have been grouped into geographic areas. We have considered here the same region repartition as in.^16^ The plots show the top 10 areas in terms of importations, along with the fluxes responsible for 60% of the importations. In the case of Europe, we show the top 20 countries, each featuring only the most significant importation flux. Fluxes between countries/areas are color-coded according to their country/area of origin.

The pattern shown here is largely consistent with the results of previous phylogeographic recon-structions .^12,14–16^ We looked more in depth to importation fluxes across European countries and found a West to East migration pattern compatible with the West-to-East gradient in peak timing observed in the region.^57,59^This highlights that human population distribution, human mobility and seasonal variation in transmission are important drivers of influenza circulation at the global and continental levels.

The multiscale nature of influenza dynamics generated by GLEAM with recurrent travel allows simultaneously reconstructing within country spread and global virus circulation, shedding light on the dynamical coupling among countries underlying seasonal epidemic waves. In Fig. 5 we compare local transmission with case importations according to the region of origin. The contribution of importations to local epidemics is important during out-of-season periods as a seeding component that can generate long transmission chains at the beginning of the influenza season. Model predictions on the geographical origin of importations may therefore carry important epidemiological information about the approaching season. Fig. 5 highlights the differences in the behavior between specific countries. A high level of geographical mixing is observed for Australia where importations during summer and at the beginning of the influenza season originate from Southeast Asia, Europe and North America. For Japan and the United States on the other hand, a geographical pattern emerges in which the large majority of importations originate from a specific region, i.e. Southeast Asia for Japan and South America for United States.

**Figure 5:**
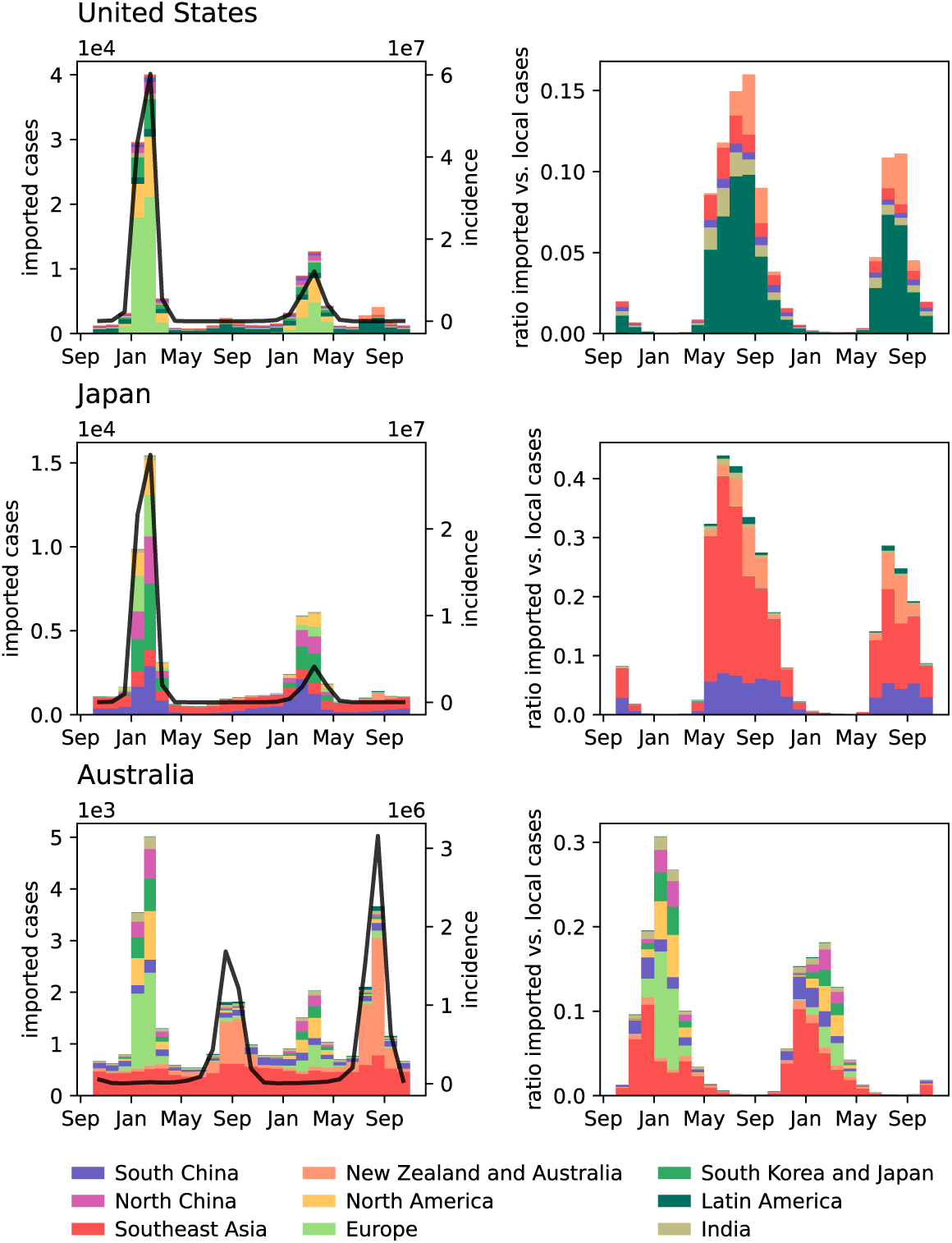
Left: simulated incidence (black line, right axis) and number of imported cases (bars, left axis) for two consecutive years from a single stochastic realization - for the best parametrization obtained for H3N2. Right: ratio of imported vs. local cases. Bars are assigned colors based on their respective are of origin (same repartition as in Fig. 4. The bars with grey/white stripes combine values for regions outside the top six from which the selected country imports cases.

### 2.4 Limitations

Our model captures global circulation patterns that largely explain both incidence and genetic data, with selected parameterizations that are generally consistent for both H3N2, H1N1 and YAM. While phylogeographic analysis showed that migration rates were substantially correlated for H3N2, H1N1, and YAM, these lineages were also characterized by different degrees of persistence in specific locations.^16^ A uniform model of seasonality in the region overlooks environmental and human forcing that are acting at the country and sub-country level and that are known to shape the epidemic dynamics in the region.^7,8,57,60^ In addition, strain-specific antigenic evolution and its interplay with demography and age structure can affect migration patterns, which is not accounted for in our model. For instance, H1N1 hits more severely the younger population that mix more at the local geographical scale but travel less frequently over long-range distances compared to the adult population.^16^ This effect has been shown to impact the spatial invasion and the local persistence of an infection.^16,61^ Differences in strain-specific patterns may arise also from complex interactions between subtypes that are difficult to capture from a general seasonal model. Further fine-tuning of simulation parameters may also assist in better capturing subtle differences in the strain specific dynamics.

## 3 Conclusions

By designing a combined approach, we were able to employ genetic data to validate and calibrate a dynamical model for the multiscale spread of influenza. The model simulates within-country epidemics, spatial coupling mediated by human mobility, and thus the resulting global circulation of the virus. The information encoded in the genetic data allowed for unambiguous identification of the essential epidemiological parameters, whereas incidence data offered only low resolution power. We were able to show that population distribution, local mobility and international travel, as well as seasonality are fundamental ingredients to accurately model influenza migration patterns.

We have here studied a decade of influenza dynamics before the COVID-19 pandemic. Following SARS-CoV-2 emergence in 2020, the global influenza circulation has been substantially altered with potential long-term consequences,^62^ as illustrated by the probable extinction of the B Yamagata variant.^63^ In such a situation, the long-term and global-scale description of influenza dynamics is more than ever important to identify viral evolutionary pathways for the prediction of vaccine composition, to inform projections on the approaching influenza season in a given region, and, on a more fundamental level, to disentangle the interplay between endogenous and exogenous factors in shaping regional epidemic waves. Phylodynamic approaches may become an invaluable tool to achieve these goals. Our study provides the starting point of a new methodological approach that can be further extended with additional ingredients and data layers to improve the description of the source-sink dynamics and strain-specific features and their interactions in the post-COVID-19 pandemic era. Advances in global influenza surveillance,^18,60^ along with the increased availability of large-scale data-sets ^8^ will undoubtedly instigate further model developments. In addition, the flexible multi-step structure of our approach makes it adaptable to a variety of epidemic models, infectious diseases, and epidemic scenarios.

## 4 Material and Methods

### 4.1 GLEAM

The GLEAM mobility layer integrates the global flight network with the daily commuting patterns between adjacent patches ^64^ (see Supplementary Material). The short-range commuting is accounted for by defining effective patch mixing, based on a time-scale separation approach.^11,28^

Air travel mobility is modeled explicitly, as a discrete-time multinomial process.^65^ In Markovian GLEAM, the daily probability that an individual travels from patch *i* to patch *j* is *p_ij_* = *w_ij_/N_i_*, with *N_i_* being the population size of *i* and *w_ij_* being the flux of passengers from *i* to *j* in the air transportation data. Traveling probability does not account for the location of residence of individuals. In the recurrent travel GLEAM, the flux of travelers *w_ij_* is subdivided into individuals resident in *i* and departing for *j*, and individuals visiting *i* and returning to the residence location *j*.^41,66–68^ Leaving and returning home are modeled as distinct processes with average trip duration assumed to be 15 days ^28^ and departing rate derived from *w_ij_* (see Supplementary Material). Influenza transmission dynamics is modeled within each patch through a compartmental model where individuals are divided in susceptible, latent, symptomatic infectious (that may or may not travel dependent on the severity of symptoms), asymptomatic infectious and recovered, i.e. immune to the virus. The average duration of the exposed and infection period are set to 1.1 and 2.5 days, respectively.^11^ Given the stochastic nature of the model, each parametrization generates a collection of possible time evolutions for the observables, such as prevalence, peak of infection, number of imported cases, etc., at the spatial resolution of a single patch and time resolution of a day, that can be aggregated at the desired level in time and space.

### Phylogeographic analysis

We combine a generalized linear model (GLM) parameterization of discrete phylogeographic diffusion ^12^ with epoch modelling^45^ in a Bayesian full probabilistic framework. Both approaches represent extensions of continuous-time Markov chain (CTMC) processes implemented in a Bayesian phylogenetic framework.^69^ The GLM diffusion model parameterises the CTMC transition rates as a log linear function of a number of potential predictors and allows estimating both the size of the contribution and the inclusion probability of each predictor. Previous applications have demonstrated how this model averaging approach can identify the predictor or set of predictors that adequately explain the dynamics among location ^12,70^ or among host transitioning. ^71^ Here, we adapt this approach to compare the fit of individual model-based fluxes as predictors of phylogeographic diffusion. In order to model heterogeneity in migration rates through time, and hence to allow for different fluxes predicting these time-variable rates, we adopt an epoch modeling approach.^45^ The epoch approach partitions evolutionary history into an arbitrary number of time intervals or epochs, separated by transition times, and allows specifying a potentially different CTMC parameterisation for each epoch. Here, we set up transition times every six months separating each time spring-summer (here defined as the period from March 21 to September 20) from autumn-winter (from the September 21 to March 20). We apply two different alternating GLM parameterizations through time: one shared by every spring-summer epoch and one shared by every autumn-winter epoch. We perform inference under the GLM and epoch model using Markov chain Monte Carlo (MCMC) integration using BEAST.^46^

We fit both time-homogeneous and epoch GLM models with the flux predictors to influenza A lineages H3N2 and H1N1 and influenza B lineages Yamagata (YAM) and Victoria (VIC), previously analysed by.^16^ These data sets consist of haemagglutinin (HA) gene sequences covering a time interval from 2000 to 2012 and represent roughly equitable spatiotemporal distributions across global regions. The data sets comprise 4,006, 2,144, 1,455, and 1,999 sequences for H3N2, H1N1, YAM, and VIC, respectively. We fit our models to the same empirical tree distributions as used in the original work. We run sufficiently long MCMC chains to ensure adequate mixing as assessed by effective sample size estimates.

## 5 Funding

PL, VC, CP, ECGB and FP acknowledge funding from EU Horizon 2020 grants MOOD (H2020-874850, publication cataloged as MOOD 102). VC acknowledges support from Horizon Europe grant ESCAPE (101095619) and Agence Nationale de la Recherche projects DATAREDUX (ANR-19-CE46-0008-03). PL, MAS and AR acknowledge funding from the European Research Council under the European Union’s Horizon 2020 research and innovation programme (grant agreement no. 725422-ReservoirDOCS) and from the Wellcome Trust Collaborative Award, 206298/Z/17/Z. MAS acknowledges support from US National Institutes of Health grants U19 AI135995, R01 AI153044 and R01 AI162611. PL acknowledges support by the Special Research Fund, KU Leuven (‘Bijzonder Onderzoeksfonds’, KU Leuven, OT/14/115), and the Research Foundation – Flanders (‘Fonds voor Wetenschappelijk Onderzoek – Vlaanderen’, G066215N, G0D5117N and G0B9317N). CP acknowledges funding by the Cariparo Foundation through the program Starting Package and the Department of Molecular Medicine of the University of Padova through the program SID from BIRD funding. TB is a Howard Hughes Medical Institute Investigator and is supported by NIH NIGMS R35 GM119774. The opinions expressed in this article are those of the authors and do not reflect the view of the National Institutes of Health, the Department of Health and Human Services, or the United States government.

## 6 Author contributions statement

T.B., V.C., C.P., P.L. designed research; F.P., E.G., T.B., V.C., C.P., P.L. performed research; F.P., E.G., T.B., M.A.S., N.S.T., A.R., V.C., C.P., P.L. analyzed data; F.P., E.G., T.B., M.A.S., N.S.T., A.R., V.C., C.P., P.L. wrote the paper.

## 7 Competing interests

The authors declare no competing interests.

## Data Availability

All data sources are available in the references of the manuscript.

https://github.com/blab/global-migration

## Notes

### Competing Interest Statement

The authors have declared no competing interest.

