## Supplementary material for "Integrating dynamical modeling and phylogeographic inference to characterize global influenza circulation": pdf

**2 Phylogeographic GLM analysis** **8**

**3 Comparative analysis of simulated fluxes** **22**

**4 Comparison simulated vs. observed epidemics** **23**

**5 Influenza migration patterns** **27**

### 1 Global Epidemic and Mobility Model

#### 1.1 Data sources

GLEAM accounts for worldwide population distribution, mobility by flight, and national commuting, modeled based on the following datasets:

- **Population distribution.** Population distribution is based on high-resolution data from the 'Gridded Population of the World' project of the Socioeconomic Data and Applications Center at Columbia University [1]. Specifically, this database provides a population estimate on a grid of cells covering the whole planet at a resolution of 15×15 minutes of arc. Those cells are assigned to nearby transportation hubs by a Voronoi-like tessellation structure that takes into account distance constraints, forming geographic census areas around each hub. The present version of the model uses 3,362 of such census areas in 220 countries.

- **Air travel.** Human mobility integrates the global flight network with the short-scale daily commuting patterns between adjacent patches. In particular, mobility by flight is modeled based on the data set provided by the International Air Transport Association and the Official Airline Guide [2] containing all worldwide origin-destination trips during 2013.

- **Commuting.** The commuting considers data from 80,000 administrative regions from 30 countries in 5 different continents [3]. Data of different spatial resolution levels were mapped into the geographical census areas formed by the Voronoi-like tessellation procedure around the main transportation hubs. The fact that census areas are relatively homogeneous allows us to estimate a gravity law that successfully reproduces the commuting data obtained across different continents and provides us with estimates for the possible commuting levels in the countries for which such data are not available [3, 4]. The mapped commuting flows can be considered as a second transport network connecting geographically close patches.

#### 1.2 Model of air-travel

##### 1.2.1 Network reconstruction and traveling process

A weighted network is built from the data by assigning to each airport pair $i$, $j$ the average daily number of passengers, $w_{ij}$, traveling between the two airports during 2013. The resulting network has $\sim 670,000$ links with a total of 7 million daily passengers. The network shows a high degree of heterogeneity both in the number of destinations per airport and in the number of passengers per connection. To avoid excessive computational times (proportional to the number of mobility links) a filtered network was generated by removing links with less than 2 passengers per day, thus keeping the high-traffic links corresponding to 16% of the total links and accounting for 98% of the total traffic. As an alternative method, we tested also the disparity filter[5] for extracting the backbone of the network - i.e. a filtering procedure that preserves the edges that represent statistically significant deviations with respect to a null model for the local assignment of weights to edges. However, simple threshold-based filtering was preferred in that, despite altering the distribution of weights, it preserved the geographic repartition of travelers.

Mobility based on air travel is modeled explicitly, as a discrete-time multinomial process with the time scale of one day [6]. Travel fluxes, $w_{ij}$, are used to compute transition probabilities that rule the travel dynamics. The recurrent and the Markovian travel approaches differ in that the former assumes individuals leave their residence patch for a certain destination and then return home after a certain time, while the second considers random traveling trajectories where the individuals have no assigned patch of residence.

##### 1.2.2 Markovian travel

Individuals in the patch $i$, belonging to the disease compartment $X_i$ (among susceptible, latent, asymptomatic infectious, etc., as described in the following section), can travel to every patch $j$ of the set $\upsilon_i$ of $i$'s neighboring populations in the air-travel network. The traveling probability from $i$ to $j$ is computed as $p_{ij} = w_{ij}/N_i$, with $N_i$ being the population of $i$. From the probability vector $\{p_{ij}\}_{j\in\upsilon_i}$ multinomial extractions yield the number of travelers $\Delta X_{ij}$ from $i$ to $j$. Once the numbers of travelers are extracted for each connection, the occupation number of the class $X_i$ in each patch is updated according to incoming and outgoing individuals

$$X_i(t) = X_i(t-1) + \sum_{j\in\upsilon_i} \Delta X_{ji} - \Delta X_{ij}.$$

##### 1.2.3 Recurrent travel

We followed the approach of [7, 8, 9, 10] here implemented in a fully stochastic individual-based version. $N_i$ individuals are assigned to the patch of residence $i$ and further subdivided into mobility classes, according to the patch where they are present at a given time. $N_{ij}(t)$ is thus the class of individuals who are resident in $i$ and traveling at time $t$ in the neighboring patch $j$, while the class $N_{ii}(t)$ groups individuals resident in $i$ and not traveling at time step $t$ – the same applies to each disease compartment $X$, in such a way that $X_{ij}(t)$ is the class of individuals in disease compartment $X$, who are resident in the patch $i$ and traveling at time $t$ in the neighboring patch $j$. At each time step, individuals of each compartment $X$ leave $i$ for the destination $j$ with probability $\nu_{ij}$ and they return to $i$ with probability $\tau^{-1}$, with $\tau$ being the average length of stay. Following these rules, mobility classes are updated at each time step with binomial and multinomial extractions.

Model parametrization is based on air-transport data and traveling statistics as follows. The length of stay on the travel destination, $\tau$, is assumed to be the same for every patch and equal to 15 days [3]. The probability of leaving per time step, $\nu_{ij}$, is instead chosen to recover the traveling fluxes of the air-travel database. To achieve this, we assume the mobility dynamics to be at equilibrium, with the occupation numbers of the mobility classes stable in time. These can be computed from the continuous differential equations

$$\partial_t N_{ii} = -\nu_i N_{ii}(t) + \tau^{-1} \sum_{j\in\upsilon_i} N_{ij}(t),$$
$$\partial_t N_{ij} = \nu_{ij} N_{ii}(t) - \tau^{-1} N_{ij}(t), \tag{1}$$

where $\upsilon_i$ is the set of $i$'s neighboring populations and $\nu_i = \sum_{j\in\upsilon_i} \nu_{ij}$. The equilibrium condition then reads

$$N_{ii} = \frac{N_i}{1+\tau\nu_i},$$
$$N_{ij} = \frac{N_i \tau \nu_{ij}}{1+\tau\nu_i}. \tag{2}$$

This implies that the flux of passengers along a connection, $w_{ij}$, can be computed as the sum of individuals leaving their residence patch and individuals returning home, namely

$$w_{ij} = \nu_{ij} N_{ii} + N_{ji}/\tau$$
$$= \frac{N_i \nu_{ij}}{1+\tau\nu_i} + \frac{N_j \nu_{ji}}{1+\tau\nu_j}. \tag{3}$$

The equilibrium condition implies the symmetry condition $w_{ij} = w_{ji}$, in that for the node population to be stable people returning home must be equal on average to people leaving. In order to compute $\nu_{ij}$ from $w_{ij}$, we note, however, that Eq. 3 provides a degenerate system of equations with the number of equations being half of the number of variables. This is due to the symmetry condition $w_{ij} = w_{ji}$, that a priori does not hold for $\nu_{ij}$. Therefore, to compute the set of leaving probabilities other constraints must be assumed. Here, we assumed that the travelers are equally subdivided into people departing and people returning home, namely $\nu_{ij} N_{ii} = N_{ji}/\tau = w_{ij}/2$. A different assumption was tested ($\nu_{ij} = \nu_{ji}$) without noticeable variation in the simulated epidemic dynamics.

##### 1.3 Model of transmission and commuting

Inside each patch of the metapopulation model, we consider a compartmental scheme, typical of influenza-like illnesses (ILIs), where each individual has a discrete disease state assigned at each moment in time among Susceptible, Latent, Asymptomatic Infectious, Symptomatic Infectious that can travel, Symptomatic Infectious that cannot travel due to the severity of symptoms, and Recovered. The rate at which a susceptible individual in

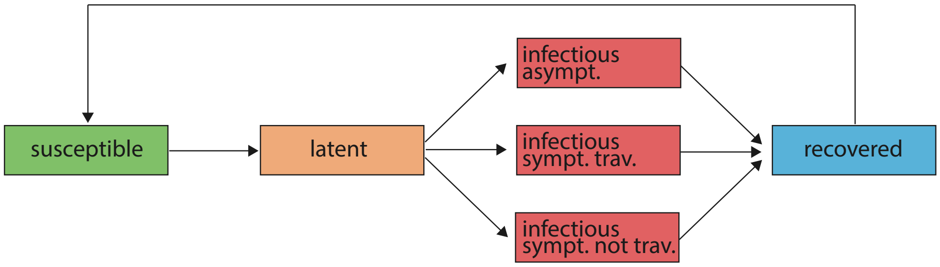

Figure 1: Scheme of the compartmental model. Expressions for the transition probabilities are reported in Tab. 2.

Table 1: Transitions between compartments and their rates.

| Transition | Type | Rate |
|---|---|---|
| $S_j \to L_j$ | Contagion | $\lambda_j$ |
| $L_j \to I_j^a$ | Spontaneous | $\varepsilon p_a$ |
| $L_j \to I_j^t$ | " | $\varepsilon(1-p_a)p_t$ |
| $L_j \to I_j^{nt}$ | " | $\varepsilon(1-p_a)(1-p_t)$ |
| $I_j^a \to R_j$ | " | $\mu$ |
| $I_j^t \to R_j$ | " | $\mu$ |
| $I_j^{nt} \to R_j$ | " | $\mu$ |
| $R_j \to S_j$ | " | $D^{-1}$ |

patch $j$ acquires the infection, the so-called force of infection $\lambda_j$, is determined by interactions with infectious persons either in the home patch $j$ or in its neighboring patches on the commuting network. Individuals mix homogeneously with other individuals sharing the same home patch $j$. The level of mixing with individuals in neighboring patches is instead modulated by commuting probability and duration, as detailed in [3, 4]. Overall transmission is modulated by the parameter $\beta_i$, representing the intrinsic viral transmissibility that depends on the patch $i$ where mixing occurs.

Given the force of infection $\lambda_j$ in patch $j$, each person in the susceptible compartment ($S_j$) contracts the infection with probability $\lambda_j \Delta t$ and enters the latent compartment ($L_j$), where $\Delta t$ is the time interval considered. Latent individuals exit the compartment with probability $\varepsilon \Delta t$, and transit to asymptomatic infectious compartment ($I_j^a$) with probability $p_a$ or, with the complementary probability $1-p_a$, become symptomatic infec-

Table 2: Summary of parameters and their values

| Parameter | Value |
|---|---|
| $\beta(t)$ | computed from Eq.( 4) |
| $\mu$ | 1/2.5 days [11] |
| $\epsilon$ | 1/1.1 days [11] |
| $p_a$ | 0.33 [12] |
| $p_t$ | 0.5 [12] |
| $r_\beta$ | 0.5 [12] |
| $D$ | explored |
| $R_{\text{min}}$ | explored |
| $R_{\text{max}}$ | explored |
| $t_{\text{max}}$ | explored |

tious. Infectious persons with symptoms are further divided between those who can travel ($I_j^t$), probability $p_t$, and those who are travel-restricted due to severe symptoms ($I_j^{nt}$) with probability $1 - p_t$. Asymptomatic individuals are assumed to have infection potential reduced by a factor $r_\beta$ with respect to individuals showing symptoms. All the infectious persons fully recover with probability $\mu\Delta t$, entering the recovered compartment ($R_j$) in the next time step. Recovered individuals lose their immunity and become fully susceptible again with rate $D^{-1}$. A schematic representation of the compartmental model is reported in Fig. 1, all transitions and corresponding rates are summarized in Table 1, while parameter values are reported in Table 2.

For the compartmental model above described the basic reproduction number ($R_0$) is given by

$$R_0 = \beta\mu^{-1}[1 - p_a + r_\beta p_a]. \tag{4}$$

##### 1.4 Seasonality in transmission

In order to model the seasonality effect in the northern and in the southern hemispheres, we follow the approach of Cooper et al. [13] rescaling the basic reproduction ratio $R_0$ by a sinusoidal function,

$$R_{0,h}(t) = \frac{R_{\text{max}}}{2}\left[\left(1 - \frac{R_{\text{min}}}{R_{\text{max}}}\right)\sin\left(\frac{2\pi}{365}(t - t_{max,h}) + \frac{\pi}{2}\right) + 1 + \frac{R_{\text{min}}}{R_{\text{max}}}\right],$$

where $h$ refers to the hemisphere considered. In the tropical region, $R_0$ is identically equal to $R_{\max}$. $t_{\max,h}$ is the time corresponding to the maximum of the sinusoid and hence to the maximum of the effective $R_0$, $R_{\max}$. The time of maximum transmissibility in the southern hemisphere, $t_{\max,s}$ occurs six months out of phase with respect to the northern hemisphere ($t_{\max,n}$). The parameters $t_{\max,n}$, $R_{\min}$ and $R_{\max}$ were calibrated as detailed in the following section. Tab. 2 summarizes all parameters and their values.

##### 1.5 Stochastic simulations

For each set of parameters and for each version of the model, we run stochastic simulations of 28-year global circulation of influenza. Simulations are in discrete time with a temporal resolution of one day. At each time step, multinomial extractions associated with traveling and infection dynamics rules are performed for each patch. To ensure that the dynamics converge to an active epidemic state, we started with an initial configuration roughly close to the equilibrium configuration. In particular, we assumed a non negligible fraction of the population to be symptomatic infectious (0.0001), a fraction $1 - 1/R_{\max}$ to be recovered, and the rest susceptible. Alternative initial conditions were tested and showed that different occupation numbers of the compartments and different distributions of infectious across patches (i.e. a single infected patch instead of all patches) were affecting the probability of observing epidemic extinction at the beginning and the intensity of the epidemic peaks during the transient stage, while, instead, the average incidence and the number of imported cases computed across years and runs, after discarding the initial epidemic period, were robust to initial conditions. The initial transient-dynamics period discarded was 8 years. We run 36 stochastic simulations for each scenario. After preliminary tests, this number was found to be sufficient to properly reconstruct the space of dynamic trajectories.

Different $R_{\max}$, $R_{\min}$, $t_{\max}$ and $D$ were compared. Specifically, values tested where: $R_{\max} \in [1.25, 1.5, 1.75, 2.0, 2.25, 2.5]$ [14]; $R_{\min} \in [0.5, 0.75]$ (exploring different levels of subcritical transmission [15]); $t_{\max} \in [Nov15th, Dec15th, Jan15th]$ and $D \in [1, 2, 4, 6, 8]$ [16, 17, 18, 19].

#### 2 Phylogeographic GLM analysis

We employ a generalized linear model (GLM) formulation of a continuous-time Markov chain (CTMC) process[20] to test how simulated fluxes predict phylogeographic diffusion. This model parameterizes the instantaneous movement rate $\Lambda_{ij}$ from location $i$ to location $j$ as a log-linear function of $P$ potential predictors $\mathbf{X}_{ij} = (x_{ij1}, \ldots, x_{ijP})'$ with unknown coefficients $\boldsymbol{\beta} = (\beta_1, \ldots, \beta_P)'$ and diagonal matrix $\boldsymbol{\delta}$ with entries $(\delta_1, \ldots, \delta_P)$. These latter unknown indicators $\delta_p \in \{0, 1\}$ determine predictor $p$'s inclusion in or exclusion from the model. We refer to [20] for the details of drawing posterior inference under the standard GLM-diffusion model. Because we aim to compare the support for individual model-based fluxes out of $P$ possible fluxes, instead of allowing all possible combinations of different predictors as in previous applications, we replace the standard transition kernel on the indicator variables that determine predictor inclusion by a new transition kernel that proposes a single randomly chosen predictor to replace the current predictor in the GLM model. The inclusion of only a single predictor removes the need to specify a prior probability over the inclusion probabilities that prefer a sparse set of predictors a priori. Instead, each flux has a prior probability of $1/P$ to be included in the model. We follow [20] in specifying that a priori all $\beta_p$ are independent and normally distributed with mean 0 and a relatively large variance of 4.

To consider time-inhomogeneity in the spatial diffusion process, we borrow epoch modeling concepts from [21]. The epoch GLM parameterizes the instantaneous movement rate $\Lambda_{ijt}$ from state $i$ to state $j$ within epoch $t$ as a log-linear function of $P$ epoch-specific predictors $\mathbf{X}_{ijt} = (x_{ijt1}, \ldots, x_{ijtP})'$ with constant-through-time, unknown coefficients $\boldsymbol{\beta}$. As epoch-specific predictors, we aggregate GLEAM fluxes for both the spring-summer and the autumn-winter periods, and we consider these as predictors for their corresponding alternating periods of time throughout the evolutionary history. We report posterior expectations for the inclusion probabilities associated with each flux.

Using this time-inhomogeneous GLM-diffusion approach, we compare fluxes based on the standard air-passenger network with Markovian and the recurrent travel version of GLEAM (Supplementary Table 3), either as seasonally aggregated fluxes over time against annually aggregated fluxes (Supplementary Table 4), as well as peak time represented by January against December(Supplementary Table 5) and November against December (Supplementary Table 6) for the full H3N2 (H3), H1N1(H1), Yamagata (YAM) and Victoria (VIC) data sets and their subsets are listed in Supplementary Tables below.

We use BEAST[22] to estimate posterior distributions for the parameters in the various phylogeographic models in conjunction with the BEAGLE library [23] to speed up likelihood calculations.

Table 3: Inclusion probabilities for the phylogeographic GLM comparison (C) of fluxes based on both Markovian (M) and recurrent travel (i.e. non-Markovian, NM) GLEAM parameterizations. The highest inclusion probability for each data set is shown in bold.

| $R_{max}$ | $R_{min}$ | D | C | $H3_{nr}$ | $H3_{r}$ | $H1_{nr}$ | $H1_{r}$ | $YAM_{nr}$ | $YAM_{r}$ | $VIC_{nr}$ | $VIC_{r}$ |
|---|---|---|---|---|---|---|---|---|---|---|---|
| (air) | (air) | (air) | (air) | 0.00 | 0.00 | 0.00 | 0.02 | 0.00 | 0.00 | 0.00 | 0.95 |
| 1.25 | 0.5 | 1 | M | 0.00 | 0.00 | 0.00 | 0.00 | 0.00 | 0.00 | 0.00 | 0.00 |
| 1.25 | 0.5 | 2 | M | 0.00 | 0.00 | 0.00 | 0.00 | 0.00 | 0.00 | 0.00 | 0.00 |
| 1.25 | 0.5 | 4 | M | 0.00 | 0.00 | 0.00 | 0.00 | 0.00 | 0.00 | 0.00 | 0.00 |
| 1.25 | 0.5 | 6 | M | 0.00 | 0.00 | 0.00 | 0.00 | 0.00 | 0.00 | 0.00 | 0.00 |
| 1.25 | 0.5 | 8 | M | 0.00 | 0.00 | 0.00 | 0.00 | 0.00 | 0.00 | 0.00 | 0.00 |
| 1.25 | 0.75 | 1 | M | 0.00 | 0.00 | 0.00 | 0.00 | 0.00 | 0.00 | 0.00 | 0.00 |
| 1.25 | 0.75 | 2 | M | 0.00 | 0.00 | 0.00 | 0.00 | 0.00 | 0.00 | 0.00 | 0.00 |
| 1.25 | 0.75 | 4 | M | 0.00 | 0.00 | 0.00 | 0.00 | 0.00 | 0.00 | 0.00 | 0.00 |
| 1.25 | 0.75 | 6 | M | 0.00 | 0.00 | 0.00 | 0.00 | 0.00 | 0.00 | 0.00 | 0.00 |
| 1.25 | 0.75 | 8 | M | 0.00 | 0.00 | 0.00 | 0.00 | 0.00 | 0.00 | 0.00 | 0.00 |
| 1.5 | 0.5 | 1 | M | 0.00 | 0.00 | 0.00 | 0.00 | 0.00 | 0.00 | 0.00 | 0.00 |
| 1.5 | 0.5 | 2 | M | 0.00 | 0.00 | 0.00 | 0.00 | 0.00 | 0.00 | 0.00 | 0.00 |
| 1.5 | 0.5 | 4 | M | 0.00 | 0.00 | 0.00 | 0.00 | 0.00 | 0.00 | 0.00 | 0.00 |
| 1.5 | 0.5 | 6 | M | 0.00 | 0.00 | 0.00 | 0.00 | 0.00 | 0.00 | 0.00 | 0.00 |
| 1.5 | 0.5 | 8 | M | 0.00 | 0.00 | 0.00 | 0.00 | 0.00 | 0.00 | 0.00 | 0.00 |
| 1.5 | 0.75 | 1 | M | 0.00 | 0.00 | 0.00 | 0.00 | 0.00 | 0.00 | 0.00 | 0.00 |
| 1.5 | 0.75 | 2 | M | 0.00 | 0.00 | 0.00 | 0.00 | 0.00 | 0.00 | 0.00 | 0.00 |
| 1.5 | 0.75 | 4 | M | 0.00 | 0.00 | 0.00 | 0.00 | 0.00 | 0.00 | 0.00 | 0.00 |
| 1.5 | 0.75 | 6 | M | 0.00 | 0.00 | 0.00 | 0.00 | 0.00 | 0.00 | 0.00 | 0.00 |
| 1.5 | 0.75 | 8 | M | 0.00 | 0.00 | 0.00 | 0.00 | 0.00 | 0.00 | 0.00 | 0.00 |
| 1.75 | 0.5 | 1 | M | 0.00 | 0.00 | 0.00 | 0.00 | 0.00 | 0.00 | 0.00 | 0.00 |
| 1.75 | 0.5 | 2 | M | 0.00 | 0.00 | 0.00 | 0.00 | 0.00 | 0.00 | 0.00 | 0.00 |
| 1.75 | 0.5 | 4 | M | 0.00 | 0.00 | 0.00 | 0.00 | 0.00 | 0.00 | 0.00 | 0.00 |
| 1.75 | 0.5 | 6 | M | 0.00 | 0.00 | 0.00 | 0.00 | 0.00 | 0.00 | 0.00 | 0.00 |
| 1.75 | 0.5 | 8 | M | 0.00 | 0.00 | 0.00 | 0.00 | 0.00 | 0.00 | 0.00 | 0.00 |
| 1.75 | 0.75 | 1 | M | 0.00 | 0.00 | 0.00 | 0.00 | 0.00 | 0.00 | 0.00 | 0.00 |
| 1.75 | 0.75 | 2 | M | 0.00 | 0.00 | 0.00 | 0.00 | 0.00 | 0.00 | 0.00 | 0.00 |
| 1.75 | 0.75 | 4 | M | 0.00 | 0.00 | 0.00 | 0.00 | 0.00 | 0.00 | 0.00 | 0.00 |
| 1.75 | 0.75 | 6 | M | 0.00 | 0.00 | 0.00 | 0.00 | 0.00 | 0.00 | 0.00 | 0.00 |
| 1.75 | 0.75 | 8 | M | 0.00 | 0.00 | 0.00 | 0.00 | 0.00 | 0.00 | 0.00 | 0.00 |
| 2 | 0.5 | 1 | M | 0.00 | 0.00 | 0.00 | 0.00 | 0.00 | 0.00 | 0.00 | 0.00 |
| 2 | 0.5 | 2 | M | 0.00 | 0.00 | 0.00 | 0.00 | 0.00 | 0.00 | 0.00 | 0.00 |
| 2 | 0.5 | 4 | M | 0.00 | 0.00 | 0.00 | 0.00 | 0.00 | 0.00 | 0.00 | 0.00 |
| 2 | 0.5 | 6 | M | 0.00 | 0.00 | 0.00 | 0.00 | 0.00 | 0.00 | 0.00 | 0.00 |

| | | | | | | | | | | | |
|---|---|---|---|---|---|---|---|---|---|---|---|
| 2 | 0.5 | 8 | M | 0.00 | 0.00 | 0.00 | 0.00 | 0.00 | 0.00 | 0.00 | 0.00 |
| 2 | 0.75 | 1 | M | 0.00 | 0.00 | 0.00 | 0.00 | 0.00 | 0.00 | 0.00 | 0.00 |
| 2 | 0.75 | 2 | M | 0.00 | 0.00 | 0.00 | 0.00 | 0.00 | 0.00 | 0.00 | 0.00 |
| 2 | 0.75 | 4 | M | 0.00 | 0.00 | 0.00 | 0.00 | 0.00 | 0.00 | 0.00 | 0.00 |
| 2 | 0.75 | 6 | M | 0.00 | 0.00 | 0.00 | 0.00 | 0.00 | 0.00 | 0.00 | 0.00 |
| 2 | 0.75 | 8 | M | 0.00 | 0.00 | 0.00 | 0.00 | 0.00 | 0.00 | 0.00 | 0.00 |
| 2.25 | 0.5 | 1 | M | 0.00 | 0.00 | 0.00 | 0.00 | 0.00 | 0.00 | 0.00 | 0.00 |
| 2.25 | 0.5 | 2 | M | 0.00 | 0.00 | 0.00 | 0.00 | 0.00 | 0.00 | 0.00 | 0.00 |
| 2.25 | 0.5 | 4 | M | 0.00 | 0.00 | 0.00 | 0.00 | 0.00 | 0.00 | 0.00 | 0.00 |
| 2.25 | 0.5 | 6 | M | 0.00 | 0.00 | 0.00 | 0.00 | 0.00 | 0.00 | 0.00 | 0.00 |
| 2.25 | 0.5 | 8 | M | 0.00 | 0.00 | 0.00 | 0.00 | 0.00 | 0.00 | 0.00 | 0.00 |
| 2.25 | 0.75 | 1 | M | 0.00 | 0.00 | 0.00 | 0.00 | 0.00 | 0.00 | 0.00 | 0.00 |
| 2.25 | 0.75 | 2 | M | 0.00 | 0.00 | 0.00 | 0.00 | 0.00 | 0.00 | 0.00 | 0.00 |
| 2.25 | 0.75 | 4 | M | 0.00 | 0.00 | 0.00 | 0.00 | 0.00 | 0.00 | 0.00 | 0.00 |
| 2.25 | 0.75 | 6 | M | 0.00 | 0.00 | 0.00 | 0.00 | 0.00 | 0.00 | 0.00 | 0.00 |
| 2.25 | 0.75 | 8 | M | 0.00 | 0.00 | 0.00 | 0.00 | 0.00 | 0.00 | 0.00 | 0.00 |
| 2.5 | 0.5 | 1 | M | 0.00 | 0.00 | 0.00 | 0.00 | 0.00 | 0.00 | 0.00 | 0.00 |
| 2.5 | 0.5 | 2 | M | 0.00 | 0.00 | 0.00 | 0.00 | 0.00 | 0.00 | 0.00 | 0.00 |
| 2.5 | 0.5 | 4 | M | 0.00 | 0.00 | 0.00 | 0.00 | 0.00 | 0.00 | 0.00 | 0.00 |
| 2.5 | 0.5 | 6 | M | 0.00 | 0.00 | 0.00 | 0.00 | 0.00 | 0.00 | 0.00 | 0.00 |
| 2.5 | 0.5 | 8 | M | 0.00 | 0.00 | 0.00 | 0.00 | 0.00 | 0.00 | 0.00 | 0.00 |
| 2.5 | 0.75 | 1 | M | 0.00 | 0.00 | 0.00 | 0.00 | 0.00 | 0.00 | 0.00 | 0.00 |
| 2.5 | 0.75 | 2 | M | 0.00 | 0.00 | 0.00 | 0.00 | 0.00 | 0.00 | 0.00 | 0.00 |
| 2.5 | 0.75 | 4 | M | 0.00 | 0.00 | 0.00 | 0.00 | 0.00 | 0.00 | 0.00 | 0.00 |
| 2.5 | 0.75 | 6 | M | 0.00 | 0.00 | 0.00 | 0.00 | 0.00 | 0.00 | 0.00 | 0.00 |
| 2.5 | 0.75 | 8 | M | 0.00 | 0.00 | 0.00 | 0.00 | 0.00 | 0.00 | 0.00 | 0.00 |
| 1.25 | 0.5 | 1 | NM | 0.00 | 0.00 | 0.00 | 0.00 | 0.00 | 0.00 | 0.00 | 0.00 |
| 1.25 | 0.5 | 2 | NM | 0.00 | 0.00 | 0.00 | 0.00 | 0.00 | 0.00 | 0.00 | 0.00 |
| 1.25 | 0.5 | 4 | NM | 0.00 | 0.00 | 0.00 | 0.00 | 0.00 | 0.00 | 0.00 | 0.00 |
| 1.25 | 0.5 | 6 | NM | 0.00 | 0.00 | 0.00 | 0.00 | 0.00 | 0.00 | 0.00 | 0.00 |
| 1.25 | 0.5 | 8 | NM | 0.00 | 0.00 | 0.00 | 0.00 | 0.00 | 0.00 | 0.00 | 0.00 |
| 1.25 | 0.75 | 1 | NM | 0.00 | 0.00 | 0.00 | 0.00 | 0.00 | 0.00 | 0.00 | 0.00 |
| 1.25 | 0.75 | 2 | NM | 0.00 | 0.00 | 0.00 | 0.00 | 0.00 | 0.00 | 0.00 | 0.00 |
| 1.25 | 0.75 | 4 | NM | 0.00 | 0.00 | 0.00 | 0.00 | 0.00 | 0.00 | 0.00 | 0.00 |
| 1.25 | 0.75 | 6 | NM | 0.00 | 0.00 | 0.00 | 0.00 | 0.00 | 0.00 | 0.00 | 0.00 |
| 1.25 | 0.75 | 8 | NM | 0.00 | 0.00 | 0.00 | 0.00 | 0.00 | 0.00 | 0.00 | 0.00 |
| 1.5 | 0.5 | 1 | NM | 0.00 | 0.00 | 0.19 | 0.02 | 0.00 | 0.00 | 0.00 | 0.00 |
| 1.5 | 0.5 | 2 | NM | 0.00 | 0.00 | 0.01 | 0.00 | 0.00 | 0.00 | 0.00 | 0.00 |
| 1.5 | 0.5 | 4 | NM | 0.00 | 0.00 | 0.00 | 0.00 | 0.00 | 0.00 | 0.00 | 0.00 |
| 1.5 | 0.5 | 6 | NM | 0.00 | 0.00 | 0.00 | 0.00 | 0.00 | 0.00 | 0.00 | 0.00 |

| | | | | | | | | | | | |
|---|---|---|---|---|---|---|---|---|---|---|---|
| 1.5 | 0.5 | 8 | NM | 0.00 | 0.00 | 0.00 | 0.00 | 0.00 | 0.00 | 0.00 | 0.00 |
| 1.5 | 0.75 | 1 | NM | 0.00 | 0.00 | 0.32 | 0.15 | 0.00 | 0.01 | 0.01 | 0.00 |
| 1.5 | 0.75 | 2 | NM | 0.00 | 0.00 | 0.01 | 0.00 | 0.00 | 0.00 | 0.00 | 0.00 |
| 1.5 | 0.75 | 4 | NM | 0.00 | 0.00 | 0.00 | 0.00 | 0.00 | 0.00 | 0.00 | 0.00 |
| 1.5 | 0.75 | 6 | NM | 0.00 | 0.00 | 0.00 | 0.00 | 0.00 | 0.00 | 0.00 | 0.00 |
| 1.5 | 0.75 | 8 | NM | 0.00 | 0.00 | 0.00 | 0.00 | 0.00 | 0.00 | 0.00 | 0.00 |
| 1.75 | 0.5 | 1 | NM | 0.00 | 0.00 | 0.00 | 0.00 | 0.00 | 0.01 | 0.00 | 0.00 |
| 1.75 | 0.5 | 2 | NM | 0.00 | 0.00 | 0.01 | 0.01 | 0.01 | 0.02 | 0.04 | 0.00 |
| 1.75 | 0.5 | 4 | NM | 0.00 | 0.00 | 0.01 | 0.00 | 0.00 | 0.00 | 0.00 | 0.00 |
| 1.75 | 0.5 | 6 | NM | 0.00 | 0.00 | 0.00 | 0.00 | 0.00 | 0.00 | 0.00 | 0.00 |
| 1.75 | 0.5 | 8 | NM | 0.00 | 0.00 | 0.00 | 0.00 | 0.00 | 0.00 | 0.00 | 0.00 |
| 1.75 | 0.75 | 1 | NM | 0.00 | 0.00 | 0.00 | 0.01 | 0.00 | 0.01 | 0.00 | 0.00 |
| 1.75 | 0.75 | 2 | NM | 0.00 | 0.00 | 0.03 | 0.02 | 0.08 | 0.09 | 0.00 | 0.00 |
| 1.75 | 0.75 | 4 | NM | 0.00 | 0.00 | 0.00 | 0.00 | 0.00 | 0.00 | 0.00 | 0.00 |
| 1.75 | 0.75 | 6 | NM | 0.00 | 0.00 | 0.00 | 0.00 | 0.00 | 0.00 | 0.00 | 0.00 |
| 1.75 | 0.75 | 8 | NM | 0.00 | 0.00 | 0.00 | 0.00 | 0.00 | 0.00 | 0.00 | 0.00 |
| 2 | 0.5 | 1 | NM | 0.00 | 0.00 | 0.00 | 0.00 | 0.00 | 0.00 | 0.00 | 0.00 |
| 2 | 0.5 | 2 | NM | 0.00 | 0.00 | 0.00 | 0.00 | 0.03 | 0.09 | 0.05 | 0.00 |
| 2 | 0.5 | 4 | NM | 0.00 | 0.00 | 0.01 | 0.01 | 0.00 | 0.00 | 0.00 | 0.00 |
| 2 | 0.5 | 6 | NM | 0.00 | 0.00 | 0.00 | 0.01 | 0.00 | 0.00 | 0.00 | 0.00 |
| 2 | 0.5 | 8 | NM | 0.00 | 0.00 | 0.00 | 0.00 | 0.00 | 0.00 | 0.00 | 0.00 |
| 2 | 0.75 | 1 | NM | 0.00 | 0.00 | 0.00 | 0.00 | 0.00 | 0.00 | 0.00 | 0.00 |
| 2 | 0.75 | 2 | NM | 0.00 | 0.00 | 0.03 | 0.02 | 0.73 | 0.46 | 0.77 | 0.00 |
| 2 | 0.75 | 4 | NM | 0.00 | 0.00 | 0.00 | 0.02 | 0.00 | 0.01 | 0.00 | 0.00 |
| 2 | 0.75 | 6 | NM | 0.00 | 0.00 | 0.00 | 0.00 | 0.00 | 0.00 | 0.00 | 0.00 |
| 2 | 0.75 | 8 | NM | 0.00 | 0.00 | 0.00 | 0.00 | 0.00 | 0.00 | 0.00 | 0.00 |
| 2.25 | 0.5 | 1 | NM | 0.00 | 0.00 | 0.00 | 0.00 | 0.00 | 0.00 | 0.00 | 0.00 |
| 2.25 | 0.5 | 2 | NM | 0.32 | 0.00 | 0.03 | 0.04 | 0.01 | 0.08 | 0.03 | 0.00 |
| 2.25 | 0.5 | 4 | NM | 0.00 | 0.00 | 0.00 | 0.02 | 0.00 | 0.02 | 0.00 | 0.00 |
| 2.25 | 0.5 | 6 | NM | 0.00 | 0.00 | 0.00 | 0.01 | 0.00 | 0.00 | 0.00 | 0.00 |
| 2.25 | 0.5 | 8 | NM | 0.00 | 0.00 | 0.00 | 0.00 | 0.00 | 0.00 | 0.00 | 0.00 |
| 2.25 | 0.75 | 1 | NM | 0.00 | 0.00 | 0.00 | 0.00 | 0.00 | 0.00 | 0.00 | 0.00 |
| 2.25 | 0.75 | 2 | NM | 0.68 | 1.00 | 0.30 | 0.51 | 0.00 | 0.00 | 0.06 | 0.00 |
| 2.25 | 0.75 | 4 | NM | 0.00 | 0.00 | 0.00 | 0.01 | 0.00 | 0.02 | 0.01 | 0.00 |
| 2.25 | 0.75 | 6 | NM | 0.00 | 0.00 | 0.00 | 0.01 | 0.00 | 0.00 | 0.00 | 0.00 |
| 2.25 | 0.75 | 8 | NM | 0.00 | 0.00 | 0.00 | 0.00 | 0.00 | 0.00 | 0.00 | 0.00 |
| 2.5 | 0.5 | 1 | NM | 0.00 | 0.00 | 0.00 | 0.00 | 0.00 | 0.00 | 0.00 | 0.00 |
| 2.5 | 0.5 | 2 | NM | 0.00 | 0.00 | 0.00 | 0.01 | 0.00 | 0.01 | 0.00 | 0.00 |
| 2.5 | 0.5 | 4 | NM | 0.00 | 0.00 | 0.00 | 0.00 | 0.03 | 0.04 | 0.01 | 0.00 |
| 2.5 | 0.5 | 6 | NM | 0.00 | 0.00 | 0.00 | 0.02 | 0.00 | 0.00 | 0.00 | 0.00 |

| | | | | | | | | | | | |
|---|---|---|---|---|---|---|---|---|---|---|---|
| 2.5 | 0.5 | 8 | NM | 0.00 | 0.00 | 0.00 | 0.00 | 0.00 | 0.00 | 0.00 | 0.00 |
| 2.5 | 0.75 | 1 | NM | 0.00 | 0.00 | 0.00 | 0.00 | 0.00 | 0.00 | 0.00 | 0.00 |
| 2.5 | 0.75 | 2 | NM | 0.00 | 0.00 | 0.00 | 0.01 | 0.00 | 0.02 | 0.00 | 0.00 |
| 2.5 | 0.75 | 4 | NM | 0.00 | 0.00 | 0.00 | 0.00 | 0.09 | 0.07 | 0.02 | 0.00 |
| 2.5 | 0.75 | 6 | NM | 0.00 | 0.00 | 0.00 | 0.03 | 0.00 | 0.01 | 0.00 | 0.00 |
| 2.5 | 0.75 | 8 | NM | 0.00 | 0.00 | 0.00 | 0.01 | 0.00 | 0.00 | 0.00 | 0.00 |

Table 4: Inclusion probabilities for the phylogeographic GLM comparison (C) of annually-aggregated (A) and seasonally-aggregated (S) fluxes based on different in a non-Markovian GLEAM parameterizations. The highest inclusion probability for each data set is shown in bold.

| $R_{max}$ | $R_{min}$ | D | C | $H3_{nr}$ | $H3_{r}$ | $H1_{nr}$ | $H1_{r}$ | $YAM_{nr}$ | $YAM_{r}$ | $VIC_{nr}$ | $VIC_{r}$ |
|---|---|---|---|---|---|---|---|---|---|---|---|
| (air) | (air) | (air) | (air) | 0.00 | 0.00 | 0.00 | 0.02 | 0.00 | 0.00 | 0.00 | 0.95 |
| 1.25 | 0.5 | 1 | A | 0.00 | 0.00 | 0.00 | 0.00 | 0.00 | 0.00 | 0.00 | 0.00 |
| 1.25 | 0.5 | 2 | A | 0.00 | 0.00 | 0.00 | 0.00 | 0.00 | 0.00 | 0.00 | 0.00 |
| 1.25 | 0.5 | 4 | A | 0.00 | 0.00 | 0.00 | 0.00 | 0.00 | 0.00 | 0.00 | 0.00 |
| 1.25 | 0.5 | 6 | A | 0.00 | 0.00 | 0.00 | 0.00 | 0.00 | 0.00 | 0.00 | 0.00 |
| 1.25 | 0.5 | 8 | A | 0.00 | 0.00 | 0.00 | 0.00 | 0.00 | 0.00 | 0.00 | 0.00 |
| 1.25 | 0.75 | 1 | A | 0.00 | 0.00 | 0.00 | 0.00 | 0.00 | 0.00 | 0.00 | 0.00 |
| 1.25 | 0.75 | 2 | A | 0.00 | 0.00 | 0.00 | 0.00 | 0.00 | 0.00 | 0.00 | 0.00 |
| 1.25 | 0.75 | 4 | A | 0.00 | 0.00 | 0.00 | 0.00 | 0.00 | 0.00 | 0.00 | 0.00 |
| 1.25 | 0.75 | 6 | A | 0.00 | 0.00 | 0.00 | 0.00 | 0.00 | 0.00 | 0.00 | 0.00 |
| 1.25 | 0.75 | 8 | A | 0.00 | 0.00 | 0.00 | 0.00 | 0.00 | 0.00 | 0.00 | 0.00 |
| 1.5 | 0.5 | 1 | A | 0.00 | 0.00 | 0.00 | 0.00 | 0.00 | 0.00 | 0.00 | 0.00 |
| 1.5 | 0.5 | 2 | A | 0.00 | 0.00 | 0.00 | 0.00 | 0.00 | 0.00 | 0.00 | 0.00 |
| 1.5 | 0.5 | 4 | A | 0.00 | 0.00 | 0.00 | 0.00 | 0.00 | 0.00 | 0.00 | 0.00 |
| 1.5 | 0.5 | 6 | A | 0.00 | 0.00 | 0.00 | 0.00 | 0.00 | 0.00 | 0.00 | 0.00 |
| 1.5 | 0.5 | 8 | A | 0.00 | 0.00 | 0.00 | 0.00 | 0.00 | 0.00 | 0.00 | 0.00 |
| 1.5 | 0.75 | 1 | A | 0.00 | 0.00 | 0.00 | 0.00 | 0.00 | 0.00 | 0.00 | 0.00 |
| 1.5 | 0.75 | 2 | A | 0.00 | 0.00 | 0.00 | 0.00 | 0.00 | 0.00 | 0.00 | 0.00 |
| 1.5 | 0.75 | 4 | A | 0.00 | 0.00 | 0.00 | 0.00 | 0.00 | 0.00 | 0.00 | 0.00 |
| 1.5 | 0.75 | 6 | A | 0.00 | 0.00 | 0.00 | 0.00 | 0.00 | 0.00 | 0.00 | 0.00 |
| 1.5 | 0.75 | 8 | A | 0.00 | 0.00 | 0.00 | 0.00 | 0.00 | 0.00 | 0.00 | 0.00 |
| 1.75 | 0.5 | 1 | A | 0.00 | 0.00 | 0.00 | 0.00 | 0.00 | 0.00 | 0.00 | 0.00 |
| 1.75 | 0.5 | 2 | A | 0.00 | 0.00 | 0.00 | 0.00 | 0.00 | 0.00 | 0.00 | 0.00 |
| 1.75 | 0.5 | 4 | A | 0.00 | 0.00 | 0.00 | 0.00 | 0.00 | 0.00 | 0.00 | 0.00 |
| 1.75 | 0.5 | 6 | A | 0.00 | 0.00 | 0.00 | 0.00 | 0.00 | 0.00 | 0.00 | 0.00 |
| 1.75 | 0.5 | 8 | A | 0.00 | 0.00 | 0.00 | 0.00 | 0.00 | 0.00 | 0.00 | 0.00 |

| | | | | | | | | | | | |
|---|---|---|---|---|---|---|---|---|---|---|---|
| 1.75 | 0.75 | 1 | A | 0.00 | 0.00 | 0.00 | 0.00 | 0.00 | 0.00 | 0.00 | 0.00 |
| 1.75 | 0.75 | 2 | A | 0.00 | 0.00 | 0.00 | 0.00 | 0.00 | 0.00 | 0.00 | 0.00 |
| 1.75 | 0.75 | 4 | A | 0.00 | 0.00 | 0.00 | 0.00 | 0.00 | 0.00 | 0.00 | 0.00 |
| 1.75 | 0.75 | 6 | A | 0.00 | 0.00 | 0.00 | 0.00 | 0.00 | 0.00 | 0.00 | 0.00 |
| 1.75 | 0.75 | 8 | A | 0.00 | 0.00 | 0.00 | 0.00 | 0.00 | 0.00 | 0.00 | 0.00 |
| 2 | 0.5 | 1 | A | 0.00 | 0.00 | 0.00 | 0.00 | 0.00 | 0.00 | 0.00 | 0.00 |
| 2 | 0.5 | 2 | A | 0.00 | 0.00 | 0.00 | 0.00 | 0.00 | 0.00 | 0.00 | 0.00 |
| 2 | 0.5 | 4 | A | 0.00 | 0.00 | 0.00 | 0.00 | 0.00 | 0.00 | 0.00 | 0.00 |
| 2 | 0.5 | 6 | A | 0.00 | 0.00 | 0.00 | 0.00 | 0.00 | 0.00 | 0.00 | 0.00 |
| 2 | 0.5 | 8 | A | 0.00 | 0.00 | 0.00 | 0.00 | 0.00 | 0.00 | 0.00 | 0.00 |
| 2 | 0.75 | 1 | A | 0.00 | 0.00 | 0.00 | 0.00 | 0.00 | 0.00 | 0.00 | 0.00 |
| 2 | 0.75 | 2 | A | 0.00 | 0.00 | 0.00 | 0.00 | 0.00 | 0.00 | 0.00 | 0.00 |
| 2 | 0.75 | 4 | A | 0.00 | 0.00 | 0.00 | 0.00 | 0.00 | 0.00 | 0.00 | 0.00 |
| 2 | 0.75 | 6 | A | 0.00 | 0.00 | 0.00 | 0.00 | 0.00 | 0.00 | 0.00 | 0.00 |
| 2 | 0.75 | 8 | A | 0.00 | 0.00 | 0.00 | 0.00 | 0.00 | 0.00 | 0.00 | 0.00 |
| 2.25 | 0.5 | 1 | A | 0.00 | 0.00 | 0.00 | 0.00 | 0.00 | 0.00 | 0.00 | 0.00 |
| 2.25 | 0.5 | 2 | A | 0.00 | 0.00 | 0.00 | 0.00 | 0.00 | 0.00 | 0.00 | 0.00 |
| 2.25 | 0.5 | 4 | A | 0.00 | 0.00 | 0.00 | 0.00 | 0.00 | 0.00 | 0.00 | 0.00 |
| 2.25 | 0.5 | 6 | A | 0.00 | 0.00 | 0.00 | 0.00 | 0.00 | 0.00 | 0.00 | 0.00 |
| 2.25 | 0.5 | 8 | A | 0.00 | 0.00 | 0.00 | 0.00 | 0.00 | 0.00 | 0.00 | 0.00 |
| 2.25 | 0.75 | 1 | A | 0.00 | 0.00 | 0.00 | 0.00 | 0.00 | 0.00 | 0.00 | 0.00 |
| 2.25 | 0.75 | 2 | A | 0.00 | 0.00 | 0.00 | 0.00 | 0.00 | 0.00 | 0.00 | 0.00 |
| 2.25 | 0.75 | 4 | A | 0.00 | 0.00 | 0.00 | 0.00 | 0.00 | 0.00 | 0.00 | 0.00 |
| 2.25 | 0.75 | 6 | A | 0.00 | 0.00 | 0.00 | 0.00 | 0.00 | 0.00 | 0.00 | 0.00 |
| 2.25 | 0.75 | 8 | A | 0.00 | 0.00 | 0.00 | 0.00 | 0.00 | 0.00 | 0.00 | 0.00 |
| 2.5 | 0.5 | 1 | A | 0.00 | 0.00 | 0.00 | 0.00 | 0.00 | 0.00 | 0.00 | 0.00 |
| 2.5 | 0.5 | 2 | A | 0.00 | 0.00 | 0.00 | 0.00 | 0.00 | 0.00 | 0.00 | 0.00 |
| 2.5 | 0.5 | 4 | A | 0.00 | 0.00 | 0.00 | 0.00 | 0.00 | 0.00 | 0.00 | 0.00 |
| 2.5 | 0.5 | 6 | A | 0.00 | 0.00 | 0.00 | 0.00 | 0.00 | 0.00 | 0.00 | 0.00 |
| 2.5 | 0.5 | 8 | A | 0.00 | 0.00 | 0.00 | 0.00 | 0.00 | 0.00 | 0.00 | 0.00 |
| 2.5 | 0.75 | 1 | A | 0.00 | 0.00 | 0.00 | 0.00 | 0.00 | 0.00 | 0.00 | 0.00 |
| 2.5 | 0.75 | 2 | A | 0.00 | 0.00 | 0.00 | 0.00 | 0.00 | 0.00 | 0.00 | 0.00 |
| 2.5 | 0.75 | 4 | A | 0.00 | 0.00 | 0.00 | 0.00 | 0.00 | 0.00 | 0.00 | 0.00 |
| 2.5 | 0.75 | 6 | A | 0.00 | 0.00 | 0.00 | 0.00 | 0.00 | 0.00 | 0.00 | 0.00 |
| 2.5 | 0.75 | 8 | A | 0.00 | 0.00 | 0.00 | 0.00 | 0.00 | 0.00 | 0.00 | 0.00 |
| 1.25 | 0.5 | 1 | S | 0.00 | 0.00 | 0.00 | 0.00 | 0.00 | 0.00 | 0.00 | 0.00 |
| 1.25 | 0.5 | 2 | S | 0.00 | 0.00 | 0.00 | 0.00 | 0.00 | 0.00 | 0.00 | 0.00 |
| 1.25 | 0.5 | 4 | S | 0.00 | 0.00 | 0.00 | 0.00 | 0.00 | 0.00 | 0.00 | 0.00 |
| 1.25 | 0.5 | 6 | S | 0.00 | 0.00 | 0.00 | 0.00 | 0.00 | 0.00 | 0.00 | 0.00 |
| 1.25 | 0.5 | 8 | S | 0.00 | 0.00 | 0.00 | 0.00 | 0.00 | 0.00 | 0.00 | 0.00 |

| | | | | | | | | | | | |
|---|---|---|---|---|---|---|---|---|---|---|---|
| 1.25 | 0.75 | 1 | S | 0.00 | 0.00 | 0.00 | 0.00 | 0.00 | 0.00 | 0.00 | 0.00 |
| 1.25 | 0.75 | 2 | S | 0.00 | 0.00 | 0.00 | 0.00 | 0.00 | 0.00 | 0.00 | 0.00 |
| 1.25 | 0.75 | 4 | S | 0.00 | 0.00 | 0.00 | 0.00 | 0.00 | 0.00 | 0.00 | 0.00 |
| 1.25 | 0.75 | 6 | S | 0.00 | 0.00 | 0.00 | 0.00 | 0.00 | 0.00 | 0.00 | 0.00 |
| 1.25 | 0.75 | 8 | S | 0.00 | 0.00 | 0.00 | 0.00 | 0.00 | 0.00 | 0.00 | 0.00 |
| 1.5 | 0.5 | 1 | S | 0.00 | 0.00 | 0.20 | 0.02 | 0.00 | 0.00 | 0.00 | 0.00 |
| 1.5 | 0.5 | 2 | S | 0.00 | 0.00 | 0.01 | 0.00 | 0.00 | 0.00 | 0.00 | 0.00 |
| 1.5 | 0.5 | 4 | S | 0.00 | 0.00 | 0.00 | 0.00 | 0.00 | 0.00 | 0.00 | 0.00 |
| 1.5 | 0.5 | 6 | S | 0.00 | 0.00 | 0.00 | 0.00 | 0.00 | 0.00 | 0.00 | 0.00 |
| 1.5 | 0.5 | 8 | S | 0.00 | 0.00 | 0.00 | 0.00 | 0.00 | 0.00 | 0.00 | 0.00 |
| 1.5 | 0.75 | 1 | S | 0.00 | 0.00 | 0.32 | 0.15 | 0.00 | 0.01 | 0.01 | 0.00 |
| 1.5 | 0.75 | 2 | S | 0.00 | 0.00 | 0.01 | 0.00 | 0.00 | 0.00 | 0.00 | 0.00 |
| 1.5 | 0.75 | 4 | S | 0.00 | 0.00 | 0.00 | 0.00 | 0.00 | 0.00 | 0.00 | 0.00 |
| 1.5 | 0.75 | 6 | S | 0.00 | 0.00 | 0.00 | 0.00 | 0.00 | 0.00 | 0.00 | 0.00 |
| 1.5 | 0.75 | 8 | S | 0.00 | 0.00 | 0.00 | 0.00 | 0.00 | 0.00 | 0.00 | 0.00 |
| 1.75 | 0.5 | 1 | S | 0.00 | 0.00 | 0.00 | 0.00 | 0.00 | 0.01 | 0.00 | 0.00 |
| 1.75 | 0.5 | 2 | S | 0.00 | 0.00 | 0.01 | 0.01 | 0.01 | 0.02 | 0.04 | 0.00 |
| 1.75 | 0.5 | 4 | S | 0.00 | 0.00 | 0.01 | 0.00 | 0.00 | 0.00 | 0.00 | 0.00 |
| 1.75 | 0.5 | 6 | S | 0.00 | 0.00 | 0.00 | 0.00 | 0.00 | 0.00 | 0.00 | 0.00 |
| 1.75 | 0.5 | 8 | S | 0.00 | 0.00 | 0.00 | 0.00 | 0.00 | 0.00 | 0.00 | 0.00 |
| 1.75 | 0.75 | 1 | S | 0.00 | 0.00 | 0.00 | 0.01 | 0.00 | 0.01 | 0.00 | 0.00 |
| 1.75 | 0.75 | 2 | S | 0.00 | 0.00 | 0.03 | 0.02 | 0.09 | 0.09 | 0.00 | 0.00 |
| 1.75 | 0.75 | 4 | S | 0.00 | 0.00 | 0.00 | 0.00 | 0.00 | 0.00 | 0.00 | 0.00 |
| 1.75 | 0.75 | 6 | S | 0.00 | 0.00 | 0.00 | 0.00 | 0.00 | 0.00 | 0.00 | 0.00 |
| 1.75 | 0.75 | 8 | S | 0.00 | 0.00 | 0.00 | 0.00 | 0.00 | 0.00 | 0.00 | 0.00 |
| 2 | 0.5 | 1 | S | 0.00 | 0.00 | 0.00 | 0.00 | 0.00 | 0.00 | 0.00 | 0.00 |
| 2 | 0.5 | 2 | S | 0.00 | 0.00 | 0.00 | 0.00 | 0.03 | 0.09 | 0.06 | 0.00 |
| 2 | 0.5 | 4 | S | 0.00 | 0.00 | 0.01 | 0.01 | 0.00 | 0.00 | 0.00 | 0.00 |
| 2 | 0.5 | 6 | S | 0.00 | 0.00 | 0.01 | 0.01 | 0.00 | 0.00 | 0.00 | 0.00 |
| 2 | 0.5 | 8 | S | 0.00 | 0.00 | 0.00 | 0.00 | 0.00 | 0.00 | 0.00 | 0.00 |
| 2 | 0.75 | 1 | S | 0.00 | 0.00 | 0.00 | 0.00 | 0.00 | 0.00 | 0.00 | 0.00 |
| 2 | 0.75 | 2 | S | 0.00 | 0.00 | 0.03 | 0.02 | 0.73 | 0.46 | 0.77 | 0.00 |
| 2 | 0.75 | 4 | S | 0.00 | 0.00 | 0.00 | 0.02 | 0.00 | 0.01 | 0.00 | 0.00 |
| 2 | 0.75 | 6 | S | 0.00 | 0.00 | 0.00 | 0.00 | 0.00 | 0.00 | 0.00 | 0.00 |
| 2 | 0.75 | 8 | S | 0.00 | 0.00 | 0.00 | 0.00 | 0.00 | 0.00 | 0.00 | 0.00 |
| 2.25 | 0.5 | 1 | S | 0.00 | 0.00 | 0.00 | 0.00 | 0.00 | 0.00 | 0.00 | 0.00 |
| 2.25 | 0.5 | 2 | S | 0.32 | 0.00 | 0.03 | 0.04 | 0.01 | 0.07 | 0.03 | 0.00 |
| 2.25 | 0.5 | 4 | S | 0.00 | 0.00 | 0.00 | 0.02 | 0.00 | 0.02 | 0.00 | 0.00 |
| 2.25 | 0.5 | 6 | S | 0.00 | 0.00 | 0.00 | 0.01 | 0.00 | 0.00 | 0.00 | 0.00 |
| 2.25 | 0.5 | 8 | S | 0.00 | 0.00 | 0.00 | 0.00 | 0.00 | 0.00 | 0.00 | 0.00 |

| | | | | | | | | | | | |
|---|---|---|---|---|---|---|---|---|---|---|---|
| 2.25 | 0.75 | 1 | S | 0.00 | 0.00 | 0.00 | 0.00 | 0.00 | 0.00 | 0.00 | 0.00 |
| 2.25 | 0.75 | 2 | S | 0.68 | 0.99 | 0.29 | 0.48 | 0.00 | 0.00 | 0.06 | 0.00 |
| 2.25 | 0.75 | 4 | S | 0.00 | 0.00 | 0.00 | 0.02 | 0.00 | 0.02 | 0.01 | 0.00 |
| 2.25 | 0.75 | 6 | S | 0.00 | 0.00 | 0.00 | 0.02 | 0.00 | 0.00 | 0.00 | 0.00 |
| 2.25 | 0.75 | 8 | S | 0.00 | 0.00 | 0.00 | 0.00 | 0.00 | 0.00 | 0.00 | 0.00 |
| 2.5 | 0.5 | 1 | S | 0.00 | 0.00 | 0.00 | 0.00 | 0.00 | 0.00 | 0.00 | 0.00 |
| 2.5 | 0.5 | 2 | S | 0.00 | 0.00 | 0.00 | 0.01 | 0.00 | 0.01 | 0.00 | 0.00 |
| 2.5 | 0.5 | 4 | S | 0.00 | 0.00 | 0.00 | 0.00 | 0.03 | 0.04 | 0.01 | 0.00 |
| 2.5 | 0.5 | 6 | S | 0.00 | 0.00 | 0.01 | 0.02 | 0.00 | 0.00 | 0.00 | 0.00 |
| 2.5 | 0.5 | 8 | S | 0.00 | 0.00 | 0.00 | 0.00 | 0.00 | 0.00 | 0.00 | 0.00 |
| 2.5 | 0.75 | 1 | S | 0.00 | 0.00 | 0.00 | 0.00 | 0.00 | 0.00 | 0.00 | 0.00 |
| 2.5 | 0.75 | 2 | S | 0.00 | 0.00 | 0.00 | 0.01 | 0.00 | 0.01 | 0.00 | 0.00 |
| 2.5 | 0.75 | 4 | S | 0.00 | 0.00 | 0.00 | 0.00 | 0.09 | 0.08 | 0.02 | 0.00 |
| 2.5 | 0.75 | 6 | S | 0.00 | 0.00 | 0.00 | 0.02 | 0.00 | 0.01 | 0.00 | 0.00 |
| 2.5 | 0.75 | 8 | S | 0.00 | 0.00 | 0.00 | 0.01 | 0.00 | 0.00 | 0.00 | 0.00 |

Table 5: Inclusion probabilities for the phylogeographic GLM comparison (C) of peak time between December-aggregated (Dec) and January-aggregated (Jan) fluxes based on a non-Markovian GLEAM parameterization. The highest inclusion probability for each data set is shown in bold.

| $R_{max}$ | $R_{min}$ | D | C | $H3_{nr}$ | $H3_{r}$ | $H1_{nr}$ | $H1_{r}$ | $YAM_{nr}$ | $YAM_{r}$ | $VIC_{nr}$ | $VIC_{r}$ |
|---|---|---|---|---|---|---|---|---|---|---|---|
| (air) | (air) | (air) | (air) | 0.00 | 0.00 | 0.00 | 0.02 | 0.00 | 0.00 | 0.00 | 0.96 |
| 1.25 | 0.5 | 1 | Dec | 0.00 | 0.00 | 0.00 | 0.00 | 0.00 | 0.00 | 0.00 | 0.00 |
| 1.25 | 0.5 | 2 | Dec | 0.00 | 0.00 | 0.00 | 0.00 | 0.00 | 0.00 | 0.00 | 0.00 |
| 1.25 | 0.5 | 4 | Dec | 0.00 | 0.00 | 0.00 | 0.00 | 0.00 | 0.00 | 0.00 | 0.00 |
| 1.25 | 0.5 | 6 | Dec | 0.00 | 0.00 | 0.00 | 0.00 | 0.00 | 0.00 | 0.00 | 0.00 |
| 1.25 | 0.5 | 8 | Dec | 0.00 | 0.00 | 0.00 | 0.00 | 0.00 | 0.00 | 0.00 | 0.00 |
| 1.25 | 0.75 | 1 | Dec | 0.00 | 0.00 | 0.00 | 0.00 | 0.00 | 0.00 | 0.00 | 0.00 |
| 1.25 | 0.75 | 2 | Dec | 0.00 | 0.00 | 0.00 | 0.00 | 0.00 | 0.00 | 0.00 | 0.00 |
| 1.25 | 0.75 | 4 | Dec | 0.00 | 0.00 | 0.00 | 0.00 | 0.00 | 0.00 | 0.00 | 0.00 |
| 1.25 | 0.75 | 6 | Dec | 0.00 | 0.00 | 0.00 | 0.00 | 0.00 | 0.00 | 0.00 | 0.00 |
| 1.25 | 0.75 | 8 | Dec | 0.00 | 0.00 | 0.00 | 0.00 | 0.00 | 0.00 | 0.00 | 0.00 |
| 1.5 | 0.5 | 1 | Dec | 0.00 | 0.00 | 0.00 | 0.00 | 0.00 | 0.00 | 0.00 | 0.00 |
| 1.5 | 0.5 | 2 | Dec | 0.00 | 0.00 | 0.00 | 0.00 | 0.00 | 0.00 | 0.00 | 0.00 |
| 1.5 | 0.5 | 4 | Dec | 0.00 | 0.00 | 0.00 | 0.00 | 0.00 | 0.00 | 0.00 | 0.00 |
| 1.5 | 0.5 | 6 | Dec | 0.00 | 0.00 | 0.00 | 0.00 | 0.00 | 0.00 | 0.00 | 0.00 |
| 1.5 | 0.5 | 8 | Dec | 0.00 | 0.00 | 0.00 | 0.00 | 0.00 | 0.00 | 0.00 | 0.00 |
| 1.5 | 0.75 | 1 | Dec | 0.00 | 0.00 | 0.00 | 0.00 | 0.00 | 0.00 | 0.00 | 0.00 |

| | | | | | | | | | | | |
|---|---|---|---|---|---|---|---|---|---|---|---|
| 1.5 | 0.75 | 2 | Dec | 0.00 | 0.00 | 0.00 | 0.00 | 0.00 | 0.00 | 0.00 | 0.00 |
| 1.5 | 0.75 | 4 | Dec | 0.00 | 0.00 | 0.00 | 0.00 | 0.00 | 0.00 | 0.00 | 0.00 |
| 1.5 | 0.75 | 6 | Dec | 0.00 | 0.00 | 0.00 | 0.00 | 0.00 | 0.00 | 0.00 | 0.00 |
| 1.5 | 0.75 | 8 | Dec | 0.00 | 0.00 | 0.00 | 0.00 | 0.00 | 0.00 | 0.00 | 0.00 |
| 1.75 | 0.5 | 1 | Dec | 0.00 | 0.00 | 0.00 | 0.00 | 0.00 | 0.00 | 0.00 | 0.00 |
| 1.75 | 0.5 | 2 | Dec | 0.00 | 0.00 | 0.00 | 0.00 | 0.00 | 0.00 | 0.00 | 0.00 |
| 1.75 | 0.5 | 4 | Dec | 0.00 | 0.00 | 0.00 | 0.00 | 0.00 | 0.00 | 0.00 | 0.00 |
| 1.75 | 0.5 | 6 | Dec | 0.00 | 0.00 | 0.00 | 0.00 | 0.00 | 0.00 | 0.00 | 0.00 |
| 1.75 | 0.5 | 8 | Dec | 0.00 | 0.00 | 0.00 | 0.00 | 0.00 | 0.00 | 0.00 | 0.00 |
| 1.75 | 0.75 | 1 | Dec | 0.00 | 0.00 | 0.00 | 0.00 | 0.00 | 0.00 | 0.00 | 0.00 |
| 1.75 | 0.75 | 2 | Dec | 0.00 | 0.00 | 0.00 | 0.00 | 0.00 | 0.00 | 0.00 | 0.00 |
| 1.75 | 0.75 | 4 | Dec | 0.00 | 0.00 | 0.00 | 0.00 | 0.00 | 0.00 | 0.00 | 0.00 |
| 1.75 | 0.75 | 6 | Dec | 0.00 | 0.00 | 0.00 | 0.00 | 0.00 | 0.00 | 0.00 | 0.00 |
| 1.75 | 0.75 | 8 | Dec | 0.00 | 0.00 | 0.00 | 0.00 | 0.00 | 0.00 | 0.00 | 0.00 |
| 2 | 0.5 | 1 | Dec | 0.00 | 0.00 | 0.00 | 0.00 | 0.00 | 0.00 | 0.00 | 0.00 |
| 2 | 0.5 | 2 | Dec | 0.00 | 0.00 | 0.00 | 0.00 | 0.00 | 0.00 | 0.00 | 0.00 |
| 2 | 0.5 | 4 | Dec | 0.00 | 0.00 | 0.00 | 0.00 | 0.00 | 0.00 | 0.00 | 0.00 |
| 2 | 0.5 | 6 | Dec | 0.00 | 0.00 | 0.00 | 0.00 | 0.00 | 0.00 | 0.00 | 0.00 |
| 2 | 0.5 | 8 | Dec | 0.00 | 0.00 | 0.00 | 0.00 | 0.00 | 0.00 | 0.00 | 0.00 |
| 2 | 0.75 | 1 | Dec | 0.00 | 0.00 | 0.00 | 0.00 | 0.00 | 0.00 | 0.00 | 0.00 |
| 2 | 0.75 | 2 | Dec | 0.00 | 0.00 | 0.00 | 0.00 | 0.00 | 0.00 | 0.00 | 0.00 |
| 2 | 0.75 | 4 | Dec | 0.00 | 0.00 | 0.00 | 0.00 | 0.00 | 0.00 | 0.00 | 0.00 |
| 2 | 0.75 | 6 | Dec | 0.00 | 0.00 | 0.00 | 0.00 | 0.00 | 0.00 | 0.00 | 0.00 |
| 2 | 0.75 | 8 | Dec | 0.00 | 0.00 | 0.00 | 0.00 | 0.00 | 0.00 | 0.00 | 0.00 |
| 2.25 | 0.5 | 1 | Dec | 0.00 | 0.00 | 0.00 | 0.00 | 0.00 | 0.00 | 0.00 | 0.00 |
| 2.25 | 0.5 | 2 | Dec | 0.00 | 0.00 | 0.00 | 0.00 | 0.00 | 0.00 | 0.00 | 0.00 |
| 2.25 | 0.5 | 4 | Dec | 0.00 | 0.00 | 0.00 | 0.00 | 0.00 | 0.00 | 0.00 | 0.00 |
| 2.25 | 0.5 | 6 | Dec | 0.00 | 0.00 | 0.00 | 0.00 | 0.00 | 0.00 | 0.00 | 0.00 |
| 2.25 | 0.5 | 8 | Dec | 0.00 | 0.00 | 0.00 | 0.00 | 0.00 | 0.00 | 0.00 | 0.00 |
| 2.25 | 0.75 | 1 | Dec | 0.00 | 0.00 | 0.00 | 0.00 | 0.00 | 0.00 | 0.00 | 0.00 |
| 2.25 | 0.75 | 2 | Dec | 0.00 | 0.00 | 0.00 | 0.00 | 0.00 | 0.00 | 0.00 | 0.00 |
| 2.25 | 0.75 | 4 | Dec | 0.00 | 0.00 | 0.00 | 0.00 | 0.00 | 0.00 | 0.00 | 0.00 |
| 2.25 | 0.75 | 6 | Dec | 0.00 | 0.00 | 0.00 | 0.00 | 0.00 | 0.00 | 0.00 | 0.00 |
| 2.25 | 0.75 | 8 | Dec | 0.00 | 0.00 | 0.00 | 0.00 | 0.00 | 0.00 | 0.00 | 0.00 |
| 2.5 | 0.5 | 1 | Dec | 0.00 | 0.00 | 0.00 | 0.00 | 0.00 | 0.00 | 0.00 | 0.00 |
| 2.5 | 0.5 | 2 | Dec | 0.00 | 0.00 | 0.00 | 0.00 | 0.00 | 0.00 | 0.00 | 0.00 |
| 2.5 | 0.5 | 4 | Dec | 0.00 | 0.00 | 0.00 | 0.00 | 0.00 | 0.00 | 0.00 | 0.00 |
| 2.5 | 0.5 | 6 | Dec | 0.00 | 0.00 | 0.00 | 0.00 | 0.00 | 0.00 | 0.00 | 0.00 |
| 2.5 | 0.5 | 8 | Dec | 0.00 | 0.00 | 0.00 | 0.00 | 0.00 | 0.00 | 0.00 | 0.00 |
| 2.5 | 0.75 | 1 | Dec | 0.00 | 0.00 | 0.00 | 0.00 | 0.00 | 0.00 | 0.00 | 0.00 |

| | | | | | | | | | | | |
|---|---|---|---|---|---|---|---|---|---|---|---|
| 2.5 | 0.75 | 2 | Dec | 0.00 | 0.00 | 0.00 | 0.00 | 0.00 | 0.00 | 0.00 | 0.00 |
| 2.5 | 0.75 | 4 | Dec | 0.00 | 0.00 | 0.00 | 0.00 | 0.00 | 0.00 | 0.00 | 0.00 |
| 2.5 | 0.75 | 6 | Dec | 0.00 | 0.00 | 0.00 | 0.00 | 0.00 | 0.00 | 0.00 | 0.00 |
| 2.5 | 0.75 | 8 | Dec | 0.00 | 0.00 | 0.00 | 0.00 | 0.00 | 0.00 | 0.00 | 0.00 |
| 1.25 | 0.5 | 1 | Jan | 0.00 | 0.00 | 0.00 | 0.00 | 0.00 | 0.00 | 0.00 | 0.00 |
| 1.25 | 0.5 | 2 | Jan | 0.00 | 0.00 | 0.00 | 0.00 | 0.00 | 0.00 | 0.00 | 0.00 |
| 1.25 | 0.5 | 4 | Jan | 0.00 | 0.00 | 0.00 | 0.00 | 0.00 | 0.00 | 0.00 | 0.00 |
| 1.25 | 0.5 | 6 | Jan | 0.00 | 0.00 | 0.00 | 0.00 | 0.00 | 0.00 | 0.00 | 0.00 |
| 1.25 | 0.5 | 8 | Jan | 0.00 | 0.00 | 0.00 | 0.00 | 0.00 | 0.00 | 0.00 | 0.00 |
| 1.25 | 0.75 | 1 | Jan | 0.00 | 0.00 | 0.00 | 0.00 | 0.00 | 0.00 | 0.00 | 0.00 |
| 1.25 | 0.75 | 2 | Jan | 0.00 | 0.00 | 0.00 | 0.00 | 0.00 | 0.00 | 0.00 | 0.00 |
| 1.25 | 0.75 | 4 | Jan | 0.00 | 0.00 | 0.00 | 0.00 | 0.00 | 0.00 | 0.00 | 0.00 |
| 1.25 | 0.75 | 6 | Jan | 0.00 | 0.00 | 0.00 | 0.00 | 0.00 | 0.00 | 0.00 | 0.00 |
| 1.25 | 0.75 | 8 | Jan | 0.00 | 0.00 | 0.00 | 0.00 | 0.00 | 0.00 | 0.00 | 0.00 |
| 1.5 | 0.5 | 1 | Jan | 0.00 | 0.00 | 0.20 | 0.02 | 0.00 | 0.00 | 0.00 | 0.00 |
| 1.5 | 0.5 | 2 | Jan | 0.00 | 0.00 | 0.01 | 0.00 | 0.00 | 0.00 | 0.00 | 0.00 |
| 1.5 | 0.5 | 4 | Jan | 0.00 | 0.00 | 0.00 | 0.00 | 0.00 | 0.00 | 0.00 | 0.00 |
| 1.5 | 0.5 | 6 | Jan | 0.00 | 0.00 | 0.00 | 0.00 | 0.00 | 0.00 | 0.00 | 0.00 |
| 1.5 | 0.5 | 8 | Jan | 0.00 | 0.00 | 0.00 | 0.00 | 0.00 | 0.00 | 0.00 | 0.00 |
| 1.5 | 0.75 | 1 | Jan | 0.00 | 0.00 | 0.32 | 0.14 | 0.00 | 0.01 | 0.01 | 0.01 |
| 1.5 | 0.75 | 2 | Jan | 0.00 | 0.00 | 0.01 | 0.00 | 0.00 | 0.00 | 0.00 | 0.00 |
| 1.5 | 0.75 | 4 | Jan | 0.00 | 0.00 | 0.00 | 0.00 | 0.00 | 0.00 | 0.00 | 0.00 |
| 1.5 | 0.75 | 6 | Jan | 0.00 | 0.00 | 0.00 | 0.00 | 0.00 | 0.00 | 0.00 | 0.00 |
| 1.5 | 0.75 | 8 | Jan | 0.00 | 0.00 | 0.00 | 0.00 | 0.00 | 0.00 | 0.00 | 0.00 |
| 1.75 | 0.5 | 1 | Jan | 0.00 | 0.00 | 0.00 | 0.00 | 0.00 | 0.00 | 0.00 | 0.00 |
| 1.75 | 0.5 | 2 | Jan | 0.00 | 0.00 | 0.01 | 0.01 | 0.01 | 0.02 | 0.04 | 0.00 |
| 1.75 | 0.5 | 4 | Jan | 0.00 | 0.00 | 0.01 | 0.00 | 0.00 | 0.00 | 0.00 | 0.00 |
| 1.75 | 0.5 | 6 | Jan | 0.00 | 0.00 | 0.00 | 0.00 | 0.00 | 0.00 | 0.00 | 0.00 |
| 1.75 | 0.5 | 8 | Jan | 0.00 | 0.00 | 0.00 | 0.00 | 0.00 | 0.00 | 0.00 | 0.00 |
| 1.75 | 0.75 | 1 | Jan | 0.00 | 0.00 | 0.00 | 0.01 | 0.00 | 0.01 | 0.00 | 0.00 |
| 1.75 | 0.75 | 2 | Jan | 0.00 | 0.00 | 0.03 | 0.02 | 0.08 | 0.09 | 0.00 | 0.00 |
| 1.75 | 0.75 | 4 | Jan | 0.00 | 0.00 | 0.00 | 0.00 | 0.00 | 0.00 | 0.00 | 0.00 |
| 1.75 | 0.75 | 6 | Jan | 0.00 | 0.00 | 0.00 | 0.00 | 0.00 | 0.00 | 0.00 | 0.00 |
| 1.75 | 0.75 | 8 | Jan | 0.00 | 0.00 | 0.00 | 0.00 | 0.00 | 0.00 | 0.00 | 0.00 |
| 2 | 0.5 | 1 | Jan | 0.00 | 0.00 | 0.00 | 0.00 | 0.00 | 0.00 | 0.00 | 0.00 |
| 2 | 0.5 | 2 | Jan | 0.00 | 0.00 | 0.00 | 0.00 | 0.04 | 0.09 | 0.06 | 0.00 |
| 2 | 0.5 | 4 | Jan | 0.00 | 0.00 | 0.01 | 0.01 | 0.00 | 0.00 | 0.00 | 0.00 |
| 2 | 0.5 | 6 | Jan | 0.00 | 0.00 | 0.00 | 0.01 | 0.00 | 0.00 | 0.00 | 0.00 |
| 2 | 0.5 | 8 | Jan | 0.00 | 0.00 | 0.00 | 0.00 | 0.00 | 0.00 | 0.00 | 0.00 |
| 2 | 0.75 | 1 | Jan | 0.00 | 0.00 | 0.00 | 0.00 | 0.00 | 0.00 | 0.00 | 0.00 |

| | | | | | | | | | | | |
|---|---|---|---|---|---|---|---|---|---|---|---|
| 2 | 0.75 | 2 | Jan | 0.00 | 0.00 | 0.03 | 0.02 | 0.73 | 0.47 | 0.76 | 0.00 |
| 2 | 0.75 | 4 | Jan | 0.00 | 0.00 | 0.00 | 0.02 | 0.00 | 0.01 | 0.00 | 0.00 |
| 2 | 0.75 | 6 | Jan | 0.00 | 0.00 | 0.00 | 0.00 | 0.00 | 0.00 | 0.00 | 0.00 |
| 2 | 0.75 | 8 | Jan | 0.00 | 0.00 | 0.00 | 0.00 | 0.00 | 0.00 | 0.00 | 0.00 |
| 2.25 | 0.5 | 1 | Jan | 0.00 | 0.00 | 0.00 | 0.00 | 0.00 | 0.00 | 0.00 | 0.00 |
| 2.25 | 0.5 | 2 | Jan | 0.32 | 0.00 | 0.03 | 0.04 | 0.02 | 0.07 | 0.02 | 0.00 |
| 2.25 | 0.5 | 4 | Jan | 0.00 | 0.00 | 0.00 | 0.02 | 0.00 | 0.02 | 0.00 | 0.00 |
| 2.25 | 0.5 | 6 | Jan | 0.00 | 0.00 | 0.00 | 0.01 | 0.00 | 0.00 | 0.00 | 0.00 |
| 2.25 | 0.5 | 8 | Jan | 0.00 | 0.00 | 0.00 | 0.00 | 0.00 | 0.00 | 0.00 | 0.00 |
| 2.25 | 0.75 | 1 | Jan | 0.00 | 0.00 | 0.00 | 0.00 | 0.00 | 0.00 | 0.00 | 0.00 |
| 2.25 | 0.75 | 2 | Jan | 0.67 | 1.00 | 0.30 | 0.51 | 0.00 | 0.00 | 0.06 | 0.01 |
| 2.25 | 0.75 | 4 | Jan | 0.00 | 0.00 | 0.00 | 0.01 | 0.00 | 0.02 | 0.01 | 0.00 |
| 2.25 | 0.75 | 6 | Jan | 0.00 | 0.00 | 0.00 | 0.02 | 0.00 | 0.00 | 0.00 | 0.00 |
| 2.25 | 0.75 | 8 | Jan | 0.00 | 0.00 | 0.00 | 0.00 | 0.00 | 0.00 | 0.00 | 0.00 |
| 2.5 | 0.5 | 1 | Jan | 0.00 | 0.00 | 0.00 | 0.00 | 0.00 | 0.00 | 0.00 | 0.00 |
| 2.5 | 0.5 | 2 | Jan | 0.00 | 0.00 | 0.00 | 0.01 | 0.00 | 0.01 | 0.00 | 0.00 |
| 2.5 | 0.5 | 4 | Jan | 0.00 | 0.00 | 0.00 | 0.00 | 0.03 | 0.04 | 0.01 | 0.00 |
| 2.5 | 0.5 | 6 | Jan | 0.00 | 0.00 | 0.01 | 0.02 | 0.00 | 0.00 | 0.00 | 0.00 |
| 2.5 | 0.5 | 8 | Jan | 0.00 | 0.00 | 0.00 | 0.00 | 0.00 | 0.00 | 0.00 | 0.00 |
| 2.5 | 0.75 | 1 | Jan | 0.00 | 0.00 | 0.00 | 0.00 | 0.00 | 0.00 | 0.00 | 0.00 |
| 2.5 | 0.75 | 2 | Jan | 0.00 | 0.00 | 0.00 | 0.01 | 0.00 | 0.02 | 0.00 | 0.00 |
| 2.5 | 0.75 | 4 | Jan | 0.00 | 0.00 | 0.00 | 0.00 | 0.09 | 0.07 | 0.02 | 0.00 |
| 2.5 | 0.75 | 6 | Jan | 0.00 | 0.00 | 0.00 | 0.03 | 0.00 | 0.01 | 0.00 | 0.00 |
| 2.5 | 0.75 | 8 | Jan | 0.00 | 0.00 | 0.00 | 0.01 | 0.00 | 0.00 | 0.00 | 0.00 |

Table 6: Inclusion probabilities for the phylogeographic GLM comparison (C) of peak time between November-aggregated (Nov) and December-aggregated (Dec) fluxes based on a non-Markovian GLEAM parameterization. The highest inclusion probability for each data set is shown in bold.

| $R_{max}$ | $R_{min}$ | D | C | $H3_{nr}$ | $H3_{r}$ | $H1_{nr}$ | $H1_{r}$ | $YAM_{nr}$ | $YAM_{r}$ | $VIC_{nr}$ | $VIC_{r}$ |
|---|---|---|---|---|---|---|---|---|---|---|---|
| (air) | (air) | (air) | (air) | 0.00 | 0.00 | 0.00 | 0.91 | 0.00 | 0.03 | 0.04 | 1.00 |
| 1.25 | 0.5 | 1 | Nov | 0.00 | 0.00 | 0.00 | 0.00 | 0.00 | 0.00 | 0.00 | 0.00 |
| 1.25 | 0.5 | 2 | Nov | 0.00 | 0.00 | 0.00 | 0.00 | 0.00 | 0.00 | 0.00 | 0.00 |
| 1.25 | 0.5 | 4 | Nov | 0.00 | 0.00 | 0.00 | 0.00 | 0.00 | 0.00 | 0.00 | 0.00 |
| 1.25 | 0.5 | 6 | Nov | 0.00 | 0.00 | 0.00 | 0.00 | 0.00 | 0.00 | 0.00 | 0.00 |
| 1.25 | 0.5 | 8 | Nov | 0.00 | 0.00 | 0.00 | 0.00 | 0.00 | 0.00 | 0.00 | 0.00 |
| 1.25 | 0.75 | 1 | Nov | 0.00 | 0.00 | 0.00 | 0.00 | 0.00 | 0.00 | 0.00 | 0.00 |
| 1.25 | 0.75 | 2 | Nov | 0.00 | 0.00 | 0.00 | 0.00 | 0.00 | 0.00 | 0.00 | 0.00 |

| | | | | | | | | | | | |
|---|---|---|---|---|---|---|---|---|---|---|---|
| 1.25 | 0.75 | 4 | Nov | 0.00 | 0.00 | 0.00 | 0.00 | 0.00 | 0.00 | 0.00 | 0.00 |
| 1.25 | 0.75 | 6 | Nov | 0.00 | 0.00 | 0.00 | 0.00 | 0.00 | 0.00 | 0.00 | 0.00 |
| 1.25 | 0.75 | 8 | Nov | 0.00 | 0.00 | 0.00 | 0.00 | 0.00 | 0.00 | 0.00 | 0.00 |
| 1.5 | 0.5 | 1 | Nov | 0.00 | 0.00 | 0.00 | 0.00 | 0.00 | 0.00 | 0.00 | 0.00 |
| 1.5 | 0.5 | 2 | Nov | 0.00 | 0.00 | 0.00 | 0.00 | 0.00 | 0.00 | 0.00 | 0.00 |
| 1.5 | 0.5 | 4 | Nov | 0.00 | 0.00 | 0.00 | 0.00 | 0.00 | 0.00 | 0.00 | 0.00 |
| 1.5 | 0.5 | 6 | Nov | 0.00 | 0.00 | 0.00 | 0.00 | 0.00 | 0.00 | 0.00 | 0.00 |
| 1.5 | 0.5 | 8 | Nov | 0.00 | 0.00 | 0.00 | 0.00 | 0.00 | 0.00 | 0.00 | 0.00 |
| 1.5 | 0.75 | 1 | Nov | 0.00 | 0.00 | 0.00 | 0.00 | 0.00 | 0.00 | 0.00 | 0.00 |
| 1.5 | 0.75 | 2 | Nov | 0.00 | 0.00 | 0.00 | 0.00 | 0.00 | 0.00 | 0.00 | 0.00 |
| 1.5 | 0.75 | 4 | Nov | 0.00 | 0.00 | 0.00 | 0.00 | 0.00 | 0.00 | 0.00 | 0.00 |
| 1.5 | 0.75 | 6 | Nov | 0.00 | 0.00 | 0.00 | 0.00 | 0.00 | 0.00 | 0.00 | 0.00 |
| 1.5 | 0.75 | 8 | Nov | 0.00 | 0.00 | 0.00 | 0.00 | 0.00 | 0.00 | 0.00 | 0.00 |
| 1.75 | 0.5 | 1 | Nov | 0.00 | 0.00 | 0.00 | 0.00 | 0.00 | 0.00 | 0.00 | 0.00 |
| 1.75 | 0.5 | 2 | Nov | 0.00 | 0.00 | 0.00 | 0.00 | 0.00 | 0.00 | 0.00 | 0.00 |
| 1.75 | 0.5 | 4 | Nov | 0.00 | 0.00 | 0.00 | 0.00 | 0.00 | 0.00 | 0.00 | 0.00 |
| 1.75 | 0.5 | 6 | Nov | 0.00 | 0.00 | 0.00 | 0.00 | 0.00 | 0.00 | 0.00 | 0.00 |
| 1.75 | 0.5 | 8 | Nov | 0.00 | 0.00 | 0.00 | 0.00 | 0.00 | 0.00 | 0.00 | 0.00 |
| 1.75 | 0.75 | 1 | Nov | 0.00 | 0.00 | 0.00 | 0.00 | 0.00 | 0.00 | 0.00 | 0.00 |
| 1.75 | 0.75 | 2 | Nov | 0.00 | 0.00 | 0.00 | 0.00 | 0.00 | 0.00 | 0.00 | 0.00 |
| 1.75 | 0.75 | 4 | Nov | 0.00 | 0.00 | 0.00 | 0.00 | 0.00 | 0.00 | 0.00 | 0.00 |
| 1.75 | 0.75 | 6 | Nov | 0.00 | 0.00 | 0.00 | 0.00 | 0.00 | 0.00 | 0.00 | 0.00 |
| 1.75 | 0.75 | 8 | Nov | 0.00 | 0.00 | 0.00 | 0.00 | 0.00 | 0.00 | 0.00 | 0.00 |
| 2 | 0.5 | 1 | Nov | 0.00 | 0.00 | 0.00 | 0.00 | 0.00 | 0.00 | 0.00 | 0.00 |
| 2 | 0.5 | 2 | Nov | 0.00 | 0.00 | 0.00 | 0.00 | 0.00 | 0.00 | 0.00 | 0.00 |
| 2 | 0.5 | 4 | Nov | 0.00 | 0.00 | 0.00 | 0.00 | 0.00 | 0.00 | 0.00 | 0.00 |
| 2 | 0.5 | 6 | Nov | 0.00 | 0.00 | 0.00 | 0.00 | 0.00 | 0.00 | 0.00 | 0.00 |
| 2 | 0.5 | 8 | Nov | 0.00 | 0.00 | 0.00 | 0.00 | 0.00 | 0.00 | 0.00 | 0.00 |
| 2 | 0.75 | 1 | Nov | 0.00 | 0.00 | 0.00 | 0.00 | 0.00 | 0.00 | 0.03 | 0.00 |
| 2 | 0.75 | 2 | Nov | 0.00 | 0.00 | 0.00 | 0.00 | 0.00 | 0.00 | 0.00 | 0.00 |
| 2 | 0.75 | 4 | Nov | 0.00 | 0.00 | 0.00 | 0.00 | 0.00 | 0.00 | 0.00 | 0.00 |
| 2 | 0.75 | 6 | Nov | 0.00 | 0.00 | 0.00 | 0.00 | 0.00 | 0.00 | 0.00 | 0.00 |
| 2 | 0.75 | 8 | Nov | 0.00 | 0.00 | 0.00 | 0.00 | 0.00 | 0.00 | 0.00 | 0.00 |
| 2.25 | 0.5 | 1 | Nov | 0.00 | 0.00 | 0.00 | 0.00 | 0.00 | 0.00 | 0.01 | 0.00 |
| 2.25 | 0.5 | 2 | Nov | 0.00 | 0.00 | 0.00 | 0.00 | 0.00 | 0.00 | 0.00 | 0.00 |
| 2.25 | 0.5 | 4 | Nov | 0.00 | 0.00 | 0.00 | 0.00 | 0.00 | 0.00 | 0.00 | 0.00 |
| 2.25 | 0.5 | 6 | Nov | 0.00 | 0.00 | 0.00 | 0.00 | 0.00 | 0.00 | 0.00 | 0.00 |
| 2.25 | 0.5 | 8 | Nov | 0.00 | 0.00 | 0.00 | 0.00 | 0.00 | 0.00 | 0.00 | 0.00 |
| 2.25 | 0.75 | 1 | Nov | 0.00 | 0.01 | 0.00 | 0.00 | 0.00 | 0.00 | 0.04 | 0.00 |
| 2.25 | 0.75 | 2 | Nov | 0.00 | 0.00 | 0.00 | 0.00 | 0.00 | 0.00 | 0.00 | 0.00 |

| | | | | | | | | | | | |
|---|---|---|---|---|---|---|---|---|---|---|---|
| 2.25 | 0.75 | 4 | Nov | 0.00 | 0.00 | 0.00 | 0.00 | 0.00 | 0.00 | 0.00 | 0.00 |
| 2.25 | 0.75 | 6 | Nov | 0.00 | 0.00 | 0.00 | 0.00 | 0.00 | 0.00 | 0.00 | 0.00 |
| 2.25 | 0.75 | 8 | Nov | 0.00 | 0.00 | 0.00 | 0.00 | 0.00 | 0.00 | 0.00 | 0.00 |
| 2.5 | 0.5 | 1 | Nov | 0.00 | 0.00 | 0.00 | 0.00 | 0.00 | 0.00 | 0.00 | 0.00 |
| 2.5 | 0.5 | 2 | Nov | 0.00 | 0.00 | 0.00 | 0.00 | 0.00 | 0.00 | 0.00 | 0.00 |
| 2.5 | 0.5 | 4 | Nov | 0.00 | 0.00 | 0.00 | 0.00 | 0.00 | 0.00 | 0.00 | 0.00 |
| 2.5 | 0.5 | 6 | Nov | 0.00 | 0.00 | 0.00 | 0.00 | 0.00 | 0.00 | 0.00 | 0.00 |
| 2.5 | 0.5 | 8 | Nov | 0.00 | 0.00 | 0.00 | 0.00 | 0.00 | 0.00 | 0.00 | 0.00 |
| 2.5 | 0.75 | 1 | Nov | 0.00 | 0.01 | 0.00 | 0.00 | 0.00 | 0.01 | 0.03 | 0.00 |
| 2.5 | 0.75 | 2 | Nov | 0.00 | 0.00 | 0.00 | 0.00 | 0.00 | 0.00 | 0.00 | 0.00 |
| 2.5 | 0.75 | 4 | Nov | 0.00 | 0.00 | 0.00 | 0.00 | 0.00 | 0.01 | 0.00 | 0.00 |
| 2.5 | 0.75 | 6 | Nov | 0.00 | 0.00 | 0.00 | 0.00 | 0.00 | 0.00 | 0.00 | 0.00 |
| 2.5 | 0.75 | 8 | Nov | 0.00 | 0.00 | 0.00 | 0.00 | 0.00 | 0.00 | 0.00 | 0.00 |
| 1.25 | 0.5 | 1 | Dec | 0.00 | 0.00 | 0.00 | 0.00 | 0.00 | 0.00 | 0.00 | 0.00 |
| 1.25 | 0.5 | 2 | Dec | 0.00 | 0.00 | 0.00 | 0.00 | 0.00 | 0.00 | 0.00 | 0.00 |
| 1.25 | 0.5 | 4 | Dec | 0.00 | 0.00 | 0.00 | 0.00 | 0.00 | 0.00 | 0.00 | 0.00 |
| 1.25 | 0.5 | 6 | Dec | 0.00 | 0.00 | 0.00 | 0.00 | 0.00 | 0.00 | 0.00 | 0.00 |
| 1.25 | 0.5 | 8 | Dec | 0.00 | 0.00 | 0.00 | 0.00 | 0.00 | 0.00 | 0.00 | 0.00 |
| 1.25 | 0.75 | 1 | Dec | 0.00 | 0.00 | 0.26 | 0.00 | 0.00 | 0.00 | 0.00 | 0.00 |
| 1.25 | 0.75 | 2 | Dec | 0.00 | 0.00 | 0.02 | 0.00 | 0.00 | 0.00 | 0.00 | 0.00 |
| 1.25 | 0.75 | 4 | Dec | 0.00 | 0.00 | 0.00 | 0.00 | 0.00 | 0.00 | 0.00 | 0.00 |
| 1.25 | 0.75 | 6 | Dec | 0.00 | 0.00 | 0.00 | 0.00 | 0.00 | 0.00 | 0.00 | 0.00 |
| 1.25 | 0.75 | 8 | Dec | 0.00 | 0.00 | 0.00 | 0.00 | 0.00 | 0.00 | 0.00 | 0.00 |
| 1.5 | 0.5 | 1 | Dec | 0.00 | 0.00 | 0.00 | 0.00 | 0.00 | 0.00 | 0.00 | 0.00 |
| 1.5 | 0.5 | 2 | Dec | 0.00 | 0.00 | 0.00 | 0.00 | 0.00 | 0.00 | 0.00 | 0.00 |
| 1.5 | 0.5 | 4 | Dec | 0.00 | 0.00 | 0.00 | 0.00 | 0.00 | 0.00 | 0.00 | 0.00 |
| 1.5 | 0.5 | 6 | Dec | 0.00 | 0.00 | 0.00 | 0.00 | 0.00 | 0.00 | 0.00 | 0.00 |
| 1.5 | 0.5 | 8 | Dec | 0.00 | 0.00 | 0.00 | 0.00 | 0.00 | 0.00 | 0.00 | 0.00 |
| 1.5 | 0.75 | 1 | Dec | 0.68 | 0.92 | 0.05 | 0.03 | 0.00 | 0.07 | 0.49 | 0.00 |
| 1.5 | 0.75 | 2 | Dec | 0.30 | 0.02 | 0.21 | 0.01 | 0.02 | 0.03 | 0.22 | 0.00 |
| 1.5 | 0.75 | 4 | Dec | 0.00 | 0.00 | 0.00 | 0.00 | 0.03 | 0.02 | 0.00 | 0.00 |
| 1.5 | 0.75 | 6 | Dec | 0.00 | 0.00 | 0.24 | 0.00 | 0.01 | 0.01 | 0.00 | 0.00 |
| 1.5 | 0.75 | 8 | Dec | 0.00 | 0.00 | 0.01 | 0.00 | 0.00 | 0.00 | 0.00 | 0.00 |
| 1.75 | 0.5 | 1 | Dec | 0.00 | 0.00 | 0.00 | 0.00 | 0.00 | 0.00 | 0.00 | 0.00 |
| 1.75 | 0.5 | 2 | Dec | 0.00 | 0.00 | 0.00 | 0.00 | 0.00 | 0.00 | 0.00 | 0.00 |
| 1.75 | 0.5 | 4 | Dec | 0.00 | 0.00 | 0.00 | 0.00 | 0.00 | 0.01 | 0.00 | 0.00 |
| 1.75 | 0.5 | 6 | Dec | 0.00 | 0.00 | 0.00 | 0.00 | 0.00 | 0.00 | 0.00 | 0.00 |
| 1.75 | 0.5 | 8 | Dec | 0.00 | 0.00 | 0.00 | 0.00 | 0.00 | 0.00 | 0.00 | 0.00 |
| 1.75 | 0.75 | 1 | Dec | 0.00 | 0.00 | 0.00 | 0.00 | 0.00 | 0.00 | 0.00 | 0.00 |
| 1.75 | 0.75 | 2 | Dec | 0.00 | 0.00 | 0.00 | 0.00 | 0.01 | 0.03 | 0.03 | 0.00 |

| | | | | | | | | | | | |
|---|---|---|---|---|---|---|---|---|---|---|---|
| 1.75 | 0.75 | 4 | Dec | 0.01 | 0.04 | 0.03 | 0.01 | 0.05 | 0.07 | 0.03 | 0.00 |
| 1.75 | 0.75 | 6 | Dec | 0.00 | 0.00 | 0.13 | 0.03 | 0.20 | 0.13 | 0.00 | 0.00 |
| 1.75 | 0.75 | 8 | Dec | 0.00 | 0.00 | 0.04 | 0.01 | 0.17 | 0.07 | 0.00 | 0.00 |
| 2 | 0.5 | 1 | Dec | 0.00 | 0.00 | 0.00 | 0.00 | 0.00 | 0.00 | 0.00 | 0.00 |
| 2 | 0.5 | 2 | Dec | 0.00 | 0.00 | 0.00 | 0.00 | 0.00 | 0.00 | 0.00 | 0.00 |
| 2 | 0.5 | 4 | Dec | 0.00 | 0.00 | 0.00 | 0.00 | 0.00 | 0.00 | 0.00 | 0.00 |
| 2 | 0.5 | 6 | Dec | 0.00 | 0.00 | 0.00 | 0.00 | 0.00 | 0.00 | 0.00 | 0.00 |
| 2 | 0.5 | 8 | Dec | 0.00 | 0.00 | 0.00 | 0.00 | 0.01 | 0.01 | 0.00 | 0.00 |
| 2 | 0.75 | 1 | Dec | 0.00 | 0.00 | 0.00 | 0.00 | 0.00 | 0.00 | 0.00 | 0.00 |
| 2 | 0.75 | 2 | Dec | 0.00 | 0.00 | 0.00 | 0.00 | 0.00 | 0.02 | 0.01 | 0.00 |
| 2 | 0.75 | 4 | Dec | 0.00 | 0.00 | 0.00 | 0.00 | 0.00 | 0.01 | 0.01 | 0.00 |
| 2 | 0.75 | 6 | Dec | 0.00 | 0.00 | 0.00 | 0.00 | 0.06 | 0.07 | 0.01 | 0.00 |
| 2 | 0.75 | 8 | Dec | 0.00 | 0.00 | 0.00 | 0.00 | 0.24 | 0.17 | 0.00 | 0.00 |
| 2.25 | 0.5 | 1 | Dec | 0.00 | 0.00 | 0.00 | 0.00 | 0.00 | 0.00 | 0.00 | 0.00 |
| 2.25 | 0.5 | 2 | Dec | 0.00 | 0.00 | 0.00 | 0.00 | 0.00 | 0.00 | 0.00 | 0.00 |
| 2.25 | 0.5 | 4 | Dec | 0.00 | 0.00 | 0.00 | 0.00 | 0.00 | 0.00 | 0.00 | 0.00 |
| 2.25 | 0.5 | 6 | Dec | 0.00 | 0.00 | 0.00 | 0.00 | 0.00 | 0.00 | 0.00 | 0.00 |
| 2.25 | 0.5 | 8 | Dec | 0.00 | 0.00 | 0.00 | 0.00 | 0.00 | 0.00 | 0.00 | 0.00 |
| 2.25 | 0.75 | 1 | Dec | 0.00 | 0.00 | 0.00 | 0.00 | 0.00 | 0.00 | 0.00 | 0.00 |
| 2.25 | 0.75 | 2 | Dec | 0.00 | 0.00 | 0.00 | 0.00 | 0.00 | 0.01 | 0.01 | 0.00 |
| 2.25 | 0.75 | 4 | Dec | 0.00 | 0.00 | 0.00 | 0.00 | 0.02 | 0.02 | 0.00 | 0.00 |
| 2.25 | 0.75 | 6 | Dec | 0.01 | 0.00 | 0.00 | 0.00 | 0.09 | 0.04 | 0.02 | 0.00 |
| 2.25 | 0.75 | 8 | Dec | 0.00 | 0.00 | 0.00 | 0.00 | 0.02 | 0.07 | 0.00 | 0.00 |
| 2.5 | 0.5 | 1 | Dec | 0.00 | 0.00 | 0.00 | 0.00 | 0.00 | 0.00 | 0.00 | 0.00 |
| 2.5 | 0.5 | 2 | Dec | 0.00 | 0.00 | 0.00 | 0.00 | 0.00 | 0.00 | 0.00 | 0.00 |
| 2.5 | 0.5 | 4 | Dec | 0.00 | 0.00 | 0.00 | 0.00 | 0.01 | 0.01 | 0.00 | 0.00 |
| 2.5 | 0.5 | 6 | Dec | 0.00 | 0.00 | 0.00 | 0.00 | 0.00 | 0.00 | 0.00 | 0.00 |
| 2.5 | 0.5 | 8 | Dec | 0.00 | 0.00 | 0.00 | 0.00 | 0.00 | 0.00 | 0.00 | 0.00 |
| 2.5 | 0.75 | 1 | Dec | 0.00 | 0.00 | 0.00 | 0.00 | 0.00 | 0.00 | 0.00 | 0.00 |
| 2.5 | 0.75 | 2 | Dec | 0.00 | 0.00 | 0.00 | 0.00 | 0.00 | 0.00 | 0.00 | 0.00 |
| 2.5 | 0.75 | 4 | Dec | 0.00 | 0.00 | 0.00 | 0.00 | 0.02 | 0.01 | 0.00 | 0.00 |
| 2.5 | 0.75 | 6 | Dec | 0.00 | 0.00 | 0.00 | 0.00 | 0.00 | 0.00 | 0.00 | 0.00 |
| 2.5 | 0.75 | 8 | Dec | 0.00 | 0.00 | 0.00 | 0.00 | 0.00 | 0.01 | 0.00 | 0.00 |

### 3 Comparative analysis of simulated fluxes

We use a multidimensional scaling analysis (MDS) to explore the differences between the simulated travel fluxes resulting from different GLEAM parameterizations. We focus on how different parameterizations create different distributions of fluxes between autumn-winter and the spring-summer periods. To quantify these differences, first, we extract, for each parameterization, the matrix containing the difference between spring-summer and autumn-winter fluxes normalized by the total annual flux. Secondly, we compute the distance between matrices by summing the absolute differences between the individual elements. Finally, the MDS allows for visualizing these distances in a two-dimensional space. In Fig. 2, we compare, for each subtype, the fluxes generated by different values of waning immunity and transmissibility peak while maintaining $R_{\max}$ constant at the value selected by the GLM for the specific subtype. For all influenza subtypes, setting the peak of transmissibility in November and December leads to distributions between spring-summer and autumn-winter fluxes that are more similar across waning of immunity values. On the other hand, when the peak of transmissibility is in January, the waning of immunity plays an important role in differentiating the flux distributions. For all influenza subtypes, the GLM consistently selects parametrizations that include the January peak and an immunity waning value that position best-supported parametrizations at one end of the data point cluster.

In Fig. 3, we compare different waning of immunity $D$ and $R_{\max}$ with peak of transmissibility for the northern hemisphere set at January 15th. Also, in this case, the parameterization selected by the GLM is the one positioned at one extreme of the cloud of points. Despite the fact that the flux distribution pattern varies in a complex way with the epidemiological parameters, the two analyses suggest that the GLM is able to capture a genuine signal in this variation.

#### 4 Comparison simulated vs. observed epidemics

We compared simulated epidemics at the level of countries with available empirical records. Data on influenza laboratory-confirmed cases were obtained by FluNet [24], a free online database maintained by WHO since 1995. We use the FluNet weekly time series data limiting our analysis until 2019 excluding the period heavily impacted by COVID-19 and excluding years 2009 and 2010 due to the H1N1 pandemic. Furthermore, we conducted subtype-specific analyses for distinct timeframes. Within influenza A lineages, we explored H3N2 from 1999 to 2019 and H1N1 from 1999 to 2008. Concerning influenza B lineages, our focus was on the period from 2011 to 2019, as limited data exists before 2011 concerning the differentiation between Victoria and Yamagata lineages. At the country level, the

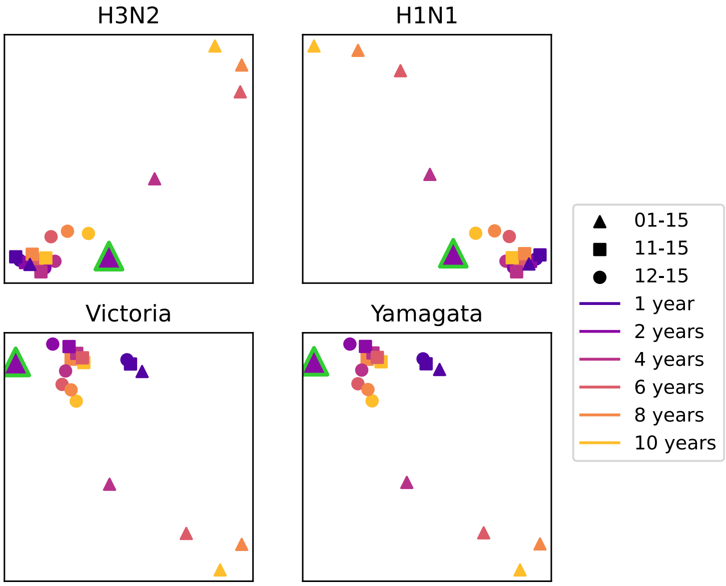

Figure 2: MDS plot illustrating flux differences for the selection of countries used for H3N2, H1N1, Yamagata, and Victoria lineages. Various symbols denote different peak transmissibility timings, while distinct colors represent the duration of waning immunity. The green-highlighted symbol depicts the parameterization with the best support. We consider the best support parametrization obtained with a predictor based on the sample size residuals for H3N2, H1N1, and Yamagata. For Victoria, we employ the analysis without the sample size residual.

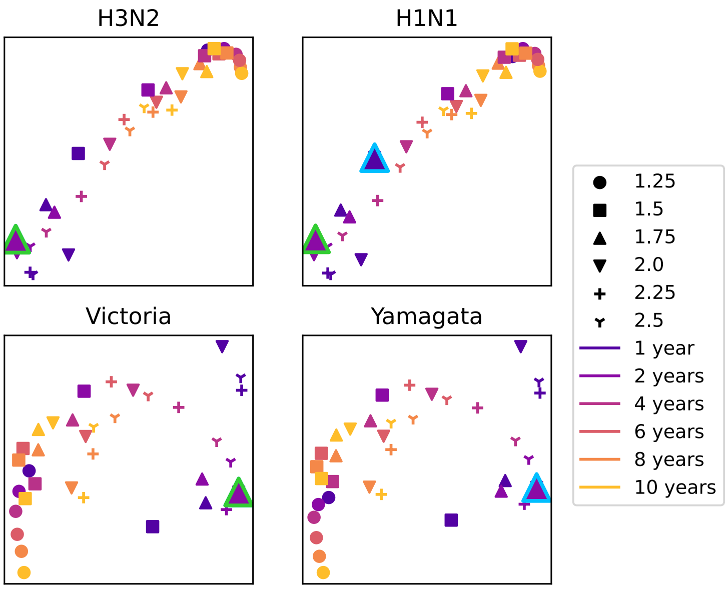

Figure 3: MDS plot illustrating flux differences for the selection of countries used for H3N2, H1N1, Yamagata, and Victoria lineages. Various symbols denote different values of $R_{\max}$, while distinct colors represent the duration of waning immunity. The green-highlighted symbol depicts parametrizations obtained with a predictor based on the sample size residuals, while the no-residual option is indicated in light blue.

Table 7: Number of countries used in the FluNet analysis, alongside the percentage relative to the total number of countries in the specified seasonal area.

| Region | H3 | H1 | YAM | VIC |
|---|---|---|---|---|
| Northern hemisphere | 64 (80%) | 36 (45%) | 34 (43%) | 28 (35%) |
| Southern hemisphere | 7 (64%) | 7 (64%) | 5 (45%) | 4 (36%) |
| Tropics | 54 (42%) | 19 (15%) | 30 (23%) | 25 (19%) |
| Worldwide | 125 (57%) | 62 (28%) | 69 (31%) | 57 (26%) |

quality and coverage of the dataset have undergone changes over time. For each country we excluded seasons with less than two months of reported cases and retained only countries with data for at least three seasons. This led to a dataset comprising 125 countries for H3N2, 62 for H1N1, 69 for Yamagata, and 57 for Victoria. For clarity, we provide separate statistics for the three seasonal areas (northern hemisphere, southern hemisphere, and tropics). A country was categorized as northern (southern) hemisphere if 75% of its urban areas were located in that hemisphere and as tropics otherwise. For all subtypes, countries considered in the analysis were distributed in the three seasonal areas. However, distribution and coverage were highly heterogeneous, as summarized in Table 7.

Further, subtyping of influenza A isolates into e.g. H3N2 or H1N1 was not available in some instances. Thus, we adjusted H3N2 totals to account for untyped A isolates, by adding each month to H3N2 counts the factor $n_A \times n_{H3N2}/(n_{H3N2} + n_{H1N1} + n_{H1N1/09} + n_{H5N1})$, where $n_A$ is the number of untyped A isolates and $n_{H3N2}$, $n_{H1N1}$, $n_{H1N1/09}$ and $n_{H5N1}$ are the number of isolates for each subtype [25]. The same was done for H1N1 and for Victoria and Yamagata by considering the untyped B data. From these records, we computed the monthly distribution of cases along the year for each year, and we averaged the profiles obtained in this way across the different seasons to obtain a single monthly distribution of incidence cases capturing robust features of the influenza epidemics of the country. We then compared these profiles with the same quantity as recovered from the simulations. We consider the best support parametrization obtained with a predictor based on the sample size residuals for H3N2, H1N1, and Yamagata. For Victoria, we employ the analysis without sample size residual, as

Table 8: Pearson correlation coefficient between FluNet and simulated epidemics. We consider the best-supported parametrizations with residual for H3N2, H1N1, and Yamagata, and the best without residual for Victoria. The table reports the average correlation values over all countries for each seasonal area and worldwide. Average correlations are computed after the Fisher transformation.

| Region | H3<br>Av. corr. | H1<br>Av. corr. | YAM<br>Av. corr. | VIC<br>Av. corr. |
|---|---|---|---|---|
| Northern hemisphere | 0.83±0.02 | 0.78±0.03 | 0.86±0.02 | 0.84±0.02 |
| Southern hemisphere | 0.66±0.08 | 0.52±0.09 | 0.79±0.03 | 0.68±0.04 |
| Tropics | 0.10±0.06 | 0.10±0.10 | 0.12±0.09 | 0.16±0.09 |
| Worldwide | 0.60±0.04 | 0.60±0.05 | 0.64±0.05 | 0.62±0.06 |

that GLEAM fluxes are not supported in analyses that include sample size residuals. Pearson correlation coefficients broken down by seasonal area are reported in Table 8.

We then tested how model performance in reproducing FluNet epidemic profiles varies across epidemiological parameters. For $R_{\min} = 0.75$ and each subtype, the bar plots of Fig. 4 summarize the average correlation, computed on over all countries varying $R_{\max}$ and $D$. The majority of scenarios tested show lower correlation compared to the best-supported parametrization. A few scenarios had a slightly higher correlation. Still, they were within the standard error of the best-supported parametrization. Therefore, the parametrization selected by the GLM is among the ones that better reproduce incidence data.

### 5 Influenza migration patterns

In Tables 10 and 9, we display the average migration fluxes among the selected regions displayed in Fig. 4, obtained from the GLEAM simulation for the two epochs. Dominant fluxes are depicted in Fig. 4 of the main paper.

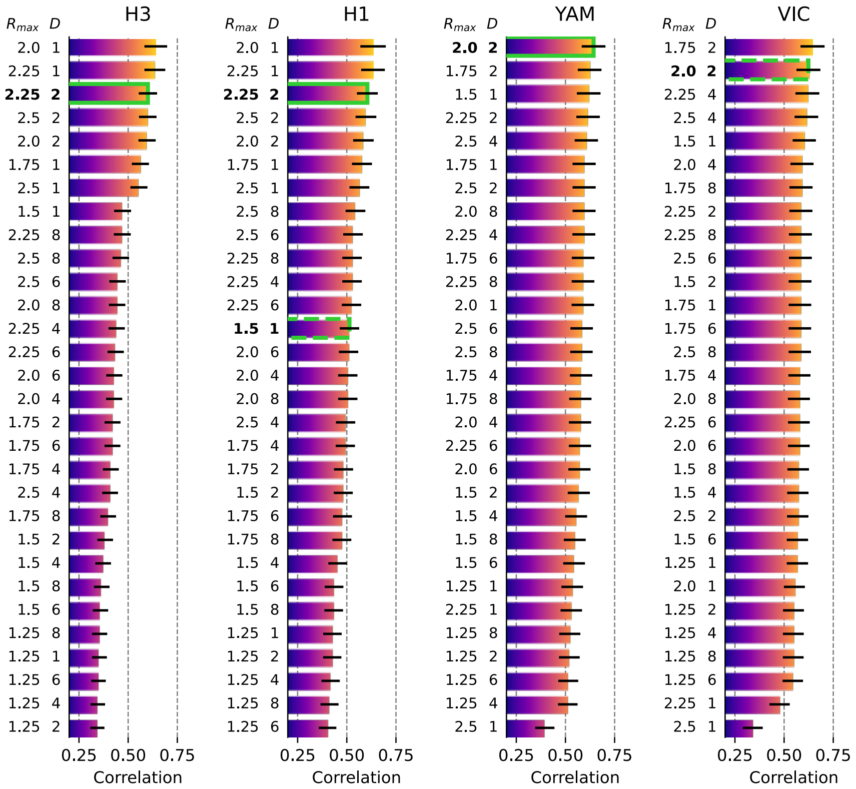

Figure 4: For each influenza subtype, the figure displays the average correlation between FluNet profiles and the simulated incidence. The parameterization receiving the highest support is highlighted in green. For H1N1, since the parametrizations with and without residuals differ, the best parametrization with residuals is highlighted with a continuous line, while the one without residual is highlighted with a dashed line. In case of Victoria, only the parametrization without residual is highlighted since, with residual no GLEaM parametrizations outperform air-travel. The error bars display the standard error.

Table 9: Simulated fluxes of imported cases among the regions used in Fig 4 of the main text, for the spring-summer period. Results for the scenario selected by GLM for H3N1, i.e. $R_{\max} = 2.25$, $R_{\min} = 0.75$ and $D = 2$ years.

| From<br><br>To | New Zealand and Australia | Latin America | India | South Korea and Japan | Europe | North China | Southeast Asia | South China | North America |
|---|---|---|---|---|---|---|---|---|---|
| New Zealand and Australia | 0 | 164 | 298 | 170 | 405 | 131 | 3274 | 565 | 252 |
| Latin America | 171 | 0 | 15 | 27 | 792 | 19 | 46 | 60 | 1149 |
| India | 638 | 32 | 0 | 53 | 346 | 31 | 2732 | 311 | 291 |
| South Korea and Japan | 1040 | 157 | 167 | 0 | 305 | 549 | 5900 | 1921 | 367 |
| Europe | 2589 | 4496 | 1261 | 325 | 0 | 236 | 3869 | 775 | 2382 |
| North China | 858 | 115 | 114 | 640 | 229 | 0 | 2814 | 22188 | 226 |
| Southeast Asia | 4870 | 64 | 2141 | 1236 | 775 | 558 | 0 | 4785 | 262 |
| South China | 1211 | 113 | 280 | 756 | 216 | 6354 | 7761 | 0 | 232 |
| North America | 1463 | 6227 | 1043 | 341 | 2039 | 192 | 1194 | 664 | 0 |

Table 10: Simulated fluxes of imported cases among the regions used in Fig 4 of the main text, for the autumn-winter period. Results for the scenario selected by GLM for H3N1, i.e. $R_{\mathrm{max}} = 2.25$, $R_{\mathrm{min}} = 0.75$ and $D = 2$ years.

| From<br>To | New Zealand and Australia | Latin America | India | South Korea and Japan | Europe | North China | Southeast Asia | South China | North America |
|---|---|---|---|---|---|---|---|---|---|
| New Zealand and Australia | 0 | 28 | 487 | 1035 | 2468 | 850 | 3172 | 1003 | 1428 |
| Latin America | 12 | 0 | 28 | 181 | 5604 | 138 | 47 | 112 | 8734 |
| India | 81 | 10 | 0 | 558 | 3659 | 356 | 3456 | 722 | 2615 |
| South Korea and Japan | 224 | 68 | 493 | 0 | 4349 | 8805 | 7924 | 8824 | 4481 |
| Europe | 395 | 2754 | 3142 | 4331 | 0 | 3539 | 5349 | 2233 | 28913 |
| North China | 131 | 53 | 310 | 8666 | 3552 | 0 | 4068 | 71975 | 2869 |
| Southeast Asia | 594 | 19 | 3650 | 10217 | 6785 | 5136 | 0 | 11799 | 2030 |
| South China | 171 | 49 | 684 | 8897 | 2335 | 78533 | 9661 | 0 | 2360 |
| North America | 237 | 4946 | 2225 | 4537 | 29456 | 2914 | 1560 | 2308 | 0 |

### References

[1] "Center for International Earth Science Information Network (CIESIN), Columbia University; and Centro Internacional de Agricultura Tropical (CIAT). The Gridded Population of the World Version 3 (GPWv3): Population Grids. Palisades, NY: Socioeconomic Data and Applications Center (SEDAC), Columbia University."

[2] "International Air Transport Association (IATA)."

[19] J. Truscott, C. Fraser, S. Cauchemez, A. Meeyai, W. Hinsley, C. A. Donnelly, A. Ghani, and N. Ferguson, "Essential epidemiological mechanisms underpinning the transmission dynamics of seasonal influenza," *Journal of The Royal Society Interface*, p. rsif20110309, June 2011.

[20] P. Lemey, A. Rambaut, T. Bedford, N. Faria, F. Bielejec, G. Baele, C. A. Russell, D. J. Smith, O. G. Pybus, D. Brockmann, and M. A. Suchard, "Unifying Viral Genetics and Human Transportation Data to Predict the Global Transmission Dynamics of Human Influenza H3n2," *PLoS Pathog*, vol. 10, p. e1003932, Feb. 2014.

[21] F. Bielejec, P. Lemey, G. Baele, A. Rambaut, and M. A. Suchard, "Inferring heterogeneous evolutionary processes through time: from sequence substitution to phylogeography," *Syst. Biol.*, vol. 63, pp. 493–504, Jul 2014.

[22] M. A. Suchard, P. Lemey, G. Baele, D. L. Ayres, A. J. Drummond, and A. Rambaut, "Bayesian phylogenetic and phylodynamic data integration using beast 1.10," *Virus Evolution*, vol. 4, no. 1, p. vey016, 2018.

[23] D. Ayres, M. Cummings, G. Baele, A. Darling, P. Lewis, J. Huelsenbeck, P. Lemey, A. Rambaut, and M. Suchard, "BEAGLE 3.0: Improved usability for a high-performance computing library for statistical phylogenetics," *Systematic Biology*, under review.

[24] N. I. C. N. of the Global Influenza Surveillance and Response System (GISRS) and World Health OrganisationWHO, "FluNet."

[25] B. S. Finkelman, C. Viboud, K. Koelle, M. J. Ferrari, N. Bharti, and B. T. Grenfell, "Global Patterns in Seasonal Activity of Influenza A/H3n2, A/H1n1, and B from 1997 to 2005: Viral Coexistence and Latitudinal Gradients," *PLoS ONE*, vol. 2, p. e1296, Dec. 2007.
